# Facilitating youth diabetes studies with the most comprehensive epidemiological dataset available through a public web portal

**DOI:** 10.1101/2023.08.02.23293517

**Authors:** Catherine McDonough, Yan Chak Li, Nita Vangeepuram, Bian Liu, Gaurav Pandey

**Affiliations:** Department of Genetics and Genomic Sciences, Icahn School of Medicine at Mount Sinai, New York, NY, USA; Department of Pediatrics, Icahn School of Medicine at Mount Sinai, New York, NY, USA; Department of Population Health Science and Policy, Icahn School of Medicine at Mount Sinai, New York, NY, USA

## Abstract

The prevalence of type 2 diabetes mellitus (DM) and prediabetes (preDM) is rapidly increasing among youth, posing significant health and economic consequences. To address this growing concern, we created the most comprehensive youth-focused diabetes dataset to date derived from National Health and Nutrition Examination Survey (NHANES) data from 1999 to 2018. The dataset, consisting of 15,149 youth aged 12 to 19 years, encompasses preDM/DM relevant variables from sociodemographic, health status, diet, and other lifestyle behavior domains. An interactive web portal, POND (Prediabetes/diabetes in youth ONline Dashboard), was developed to provide public access to the dataset, allowing users to explore variables potentially associated with youth preDM/DM. Leveraging statistical and machine learning methods, we conducted two case studies, revealing established and lesser-known variables linked to youth preDM/DM. This dataset and portal can facilitate future studies to inform prevention and management strategies for youth prediabetes and diabetes.

## Introduction

Type 2 diabetes mellitus (DM) is a complex disease influenced by several biological and epidemiological factors (*1, 2*), such as obesity (*3*), family history (*4*), diet (*1, 5*), physical activity level (*1, 6*), and socioeconomic status (*7, 8*). Prediabetes, characterized by elevated blood glucose levels below the diabetes threshold, is a precursor condition to diabetes (*9*). There has been an alarming increasing trend in the prevalence of youth with prediabetes and DM (preDM/DM) both in the United States (*10*–*16*) and worldwide (*17, 18*), and the numbers of newly diagnosed youth living with preDM/DM are also expected to increase (*10, 17, 19*). The latest estimate based on nationally representative data showed that the prevalence of preDM/DM among youth increased from 11.6% in 1999-2002 to 28.2% in 2015-2018 in the United States (*20*). This growth is particularly concerning because preDM/DM disproportionately affects racial and ethnic minority groups and those with low socioeconomic status (*7, 8, 19, 21*–*23*), leading to significant health disparities. Having preDM/DM at a younger age also confers a higher health and economic burden resulting from living with the condition for more years and a higher risk of developing other cardiometabolic diseases (*14, 24*–*28*). This serious challenge calls for increased research into factors associated with preDM/DM among youth and how they can collectively affect disease risk and inform prevention strategies.

In particular, the most critically needed research is exploring the collective impact of various risk factors across multiple health-related domains. While clinical factors, such as obesity, have been mechanistically linked to insulin resistance (*29*), it is important to consider the broader perspective. There is an increasing recognition that social determinants of health (SDoH) play a significant role in amplifying the risk of diabetes and diabetes-related disparities. For example, factors such as limited access to healthcare, food and housing insecurity, and the neighborhood-built environment have been identified as influential contributors (*7, 8, 21, 30*). However, to gain a comprehensive understanding, it is essential to delve into other less studied variables, such as screen time, acculturation, or frequency of eating out, and examine how they interact to increase the risk of preDM/DM among youth (*2*).

One of the major challenges that has limited research into youth preDM/DM risk factors is that there are no publicly available, easily accessible data comprehensively profiling interrelated epidemiological factors for young individuals (*2*). Specifically, most available public diabetes data portals focus on providing aggregated descriptive trends, such as preDM/DM prevalence for the entire population or subgroups stratified by race and ethnicity (*31*–*35*), which does not allow in-depth examination of the relationships between multiple risk factors and preDM/DM risk using individual level data. While there do exist a few individual-level public diabetes datasets (*36*–*40*), they include mainly clinical measurements, while other important risk factors such as those related to diet, physical activity, and SDoH are limited. In addition, these datasets are not available for youth populations, as they either focus exclusively on adult populations and not on youth specifically (*36, 38*–*40*). Furthermore, these datasets are not accompanied by any user-friendly online portals that can help explore or analyze these data to reveal interesting knowledge about youth preDM/DM. This shows that there is a lack of a comprehensive dataset that includes multiple epidemiological variables to study youth preDM/DM, and easily usable functionalities to explore and analyze data.

To directly address this data gap, we turned to the National Health and Nutrition Examination Survey (NHANES), which offers a promising path for examining preDM/DM among the US youth population by providing a rich source of individual- and household-level epidemiological factors. As a result, NHANES has been a prominent data source for studying youth preDM/DM trends and associated factors (*15, 41*–*44*). However, the utilization of NHANES data requires extensive data processing that is laborious and time-intensive (*45*). This represents a major challenge for the wide-spread use of these high-quality and extensive data for studying youth preDM/DM.

In this work, we directly addressed the above challenges by processing NHANES data from 1999 to 2018 into a large-scale youth diabetes-focused dataset that covers a variety of relevant variable domains, namely sociodemographic factors, health status indicators, diet and other lifestyle behaviors. We also provided public access to this high-quality comprehensive youth preDM/DM dataset, as well as functionalities to explore and analyze it, through the user-friendly Prediabetes/diabetes in youth ONline Dashboard (POND, https://rstudio-connect.hpc.mssm.edu/POND/). We demonstrated the dataset’s utility and potential through two case studies that employed statistical analyses and machine learning (ML) approaches, respectively, to identify a variety of epidemiological factors associated with youth preDM/DM. Through this work, we aimed to enable researchers to investigate the multifactorial variables associated with youth preDM/DM, which may drive advancements in prevention and management strategies.

## Results

**Fig. 1** shows the workflow of this study, including the processing of NHANES data, the development of POND, and the case studies we conducted.

**Fig. 1.**
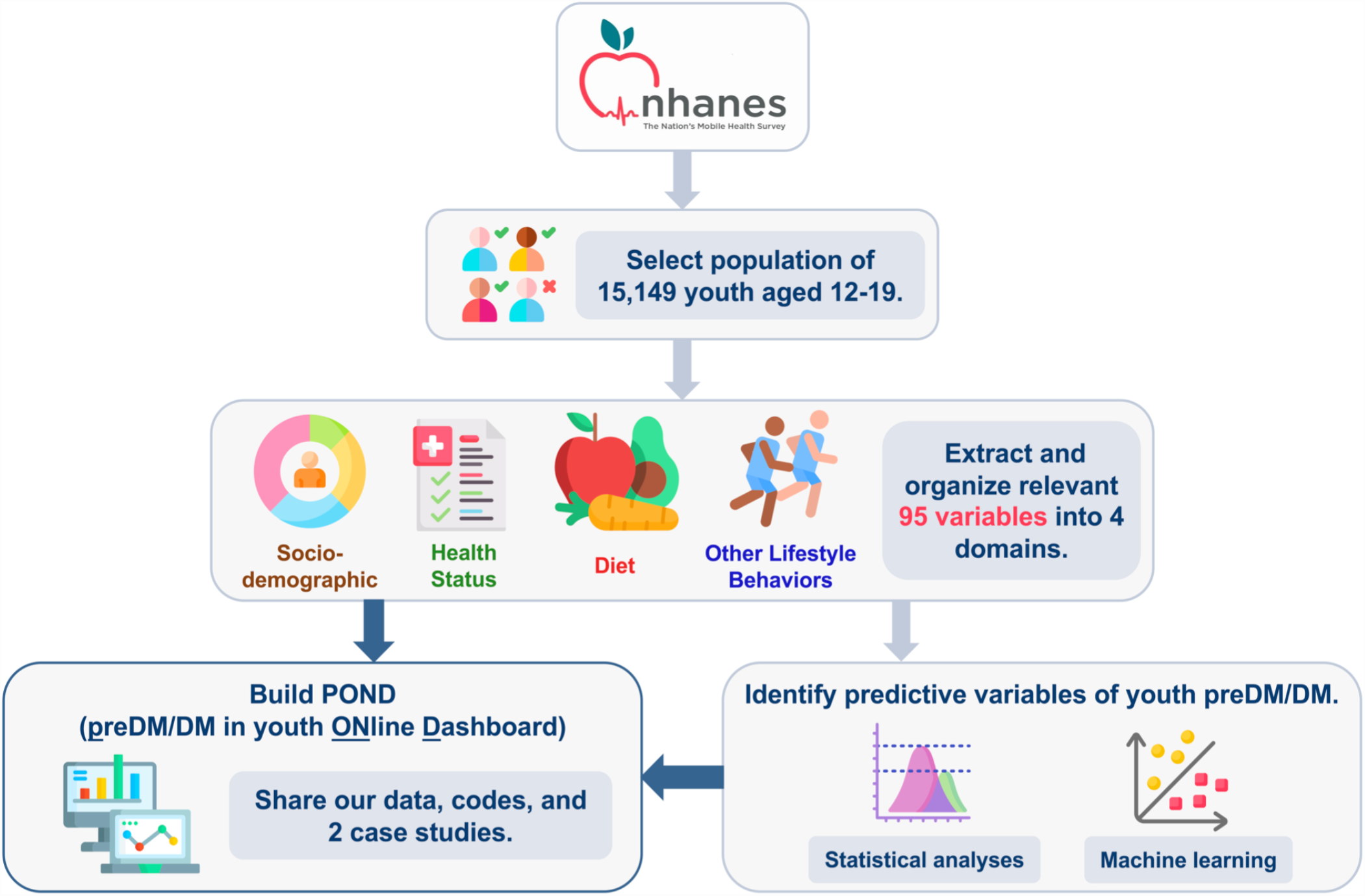
Study design and workflow. We processed data from 10 survey cycles (1999-2018) from the National Health and Nutrition Examination Survey (NHANES), which yielded 15,149 youth with known prediabetes/diabetes (preDM/DM) status. We extracted 95 variables that were relevant to preDM/DM and organized them into 4 domains: sociodemographic, health status, diet, and other lifestyle behaviors. We made the dataset easily accessible to the public through the user-friendly POND (Prediabetes/diabetes in youth ONline Dashboard) web portal, enabling users to navigate, visualize, and download the data. Additionally, we provided two case studies with complementary statistical and machine learning methods. Both analyses identified predictive variables associated with youth diabetes, and the results can be explored in POND. (Some images in this figure were obtained from the open-source collection at https://www.flaticon.com and were made by Freepik.)

### Youth preDM/DM-focused dataset

Our study population consists of 15,149 youth aged 12 to 19 years who participated in the 1999-2018 NHANES cycles and met our selection criteria **(Fig. 2)**. Approximately 13.2% of US youth were at risk of preDM/DM according to the clinically standard criteria for defining preDM/DM per the American Diabetes Association (ADA) guidelines (fasting plasma glucose (FPG)≥100 mg/dL and/or hemoglobin A1c (HbA1c)≥5.7%) **(Table 1)**. The survey-weighted prevalence of preDM/DM in US youth rose substantially from 4.1% in 1999 to 22.0% in 2018 **(Fig. S1)**.

**Table 1.** Unweighted study population characteristics. Unweighted statistics of some key variables describing the study population in the youth preDM/DM dataset overall and by preDM/DM status. More detailed statistics for all the variables in our dataset can be found in the Data Exploration section of POND.

| Variables | Overall | With preDM/DM | without preDM/DM |
| --- | --- | --- | --- |
|  | (n=15,149) | (n=2,010)<br>(Unweighted % = 13.3) | (n=13,139) |
| *n (%) or median (interquartile range)* |  |  |  |
| **Sociodemographic** |  |  |  |
| Age (years) | 15 (13, 17) | 15 (13, 17) | 16 (14, 17) |
| Female Sex | 7430 (49.0) | 691 (34.4) | 6739 (51.3) |
| Race/ethnicity |  |  |  |
| Hispanic | 5565 (36.7) | 711 (35.4) | 4854 (36.9) |
| White, Non-Hispanic | 4033 (26.6) | 431 (21.4) | 3602 (27.4) |
| Black, Non-Hispanic | 4292 (28.3) | 676 (33.6) | 3616 (27.5) |
| Other | 1259 (8.3) | 192 (9.6) | 1067 (8.1) |
| Insurance |  |  |  |
| Private | 6392 (43.0) | 744 (37.7) | 5648 (43.8) |
| Medicare, government, or single Service | 2026 (13.6) | 268 (13.6) | 1758 (13.6) |
| Medicaid/CHIP* | 3637 (24.4) | 564 (28.6) | 3073 (23.8) |
| No insurance | 2821 (19.0) | 395 (20.0) | 2426 (18.8) |
| Authorized for food stamps | 7833 (69.4) | 1037 (61.1) | 6796 (70.8) |
| **Health status** |  |  |  |
| Body Mass Index (BMI) percentile |  |  |  |
| Underweight<br>(BMI %ile < 5th) | 462 (3.1) | 40 (2.0) | 422 (3.2) |
| Normal weight<br>(5th ≤ BMI %ile < 85th) | 8516 (56.8) | 933 (46.8) | 7583 (58.4) |
| Overweight<br>(85th ≤ BMI %ile < 95th) | 2788 (18.6) | 356 (17.9) | 2432 (18.7) |
| Obese<br>(95th ≤ BMI %ile) | 3214 (21.5) | 663 (33.3) | 2551 (19.6) |
| Hypertensive† | 2552 (17.4) | 502 (26.1) | 2050 (16.1) |
| High total cholesterol (≥ 170 mg/dL) | 4951 (33.2) | 707 (35.6) | 4244 (32.8) |
| Fasting plasma glucose (mg/dL) | 93 (88, 98) | 102 (100, 106) | 91 (86, 95) |
| Hemoglobin A1c (%) | 5.2 (5.0, 5.4) | 5.5 (5.2, 5.7) | 5.2 (5.0, 5.3) |
| **Diet** |  |  |  |
| Meals eaten out per week | 2 (1, 3) | 2 (1, 3) | 2 (1, 3) |
| Total grain (oz eq.) intake 24 hours prior | 6.55 (4.24, 9.66) | 6.43 (4.19, 9.58) | 6.57 (4.25, 9.67) |
| Total fruits (cup eq.) intake 24 hours prior | 0.38 (0.00, 1.44) | 0.26 (0.00, 1.37) | 0.40 (0.00, 1.45) |
| Total vegetable (cup eq.) intake 24 hours prior | 0.88 (0.39, 1.58) | 0.84 (0.37, 1.54) | 0.89 (0.39, 1.59) |
| Total protein (oz eq.) intake 24 hours prior | 5.29 (2.71, 9.15) | 4.73 (2.46, 8.37) | 5.38 (2.76, 9.34) |
| Added sugar (tsp eq.) intake 24 hours prior | 20.42 (11.49, 32.49) | 20.09 (11.15, 31.89) | 20.48 (11.57, 32.59) |
| **Other lifestyle behavior** |  |  |  |
| Physical activity minutes per week | 209 (45, 488) | 210 (49, 476) | 209 (45, 491) |
| Screen time hours per day | 5 (3, 8) | 5 (3, 8) | 5 (2, 7) |
| Exposed to secondhand smoke at home | 3297 (21.9) | 469 (23.6) | 2828 (21.7) |
| *CHIP = Child Health Insurance Program. |  |  |  |
| †Hypertensive was defined by blood pressure ≥ 90th percentile or ≥ 120/80 mm Hg for children ≥ 13 years (2). |  |  |  |

**Fig. 2.**
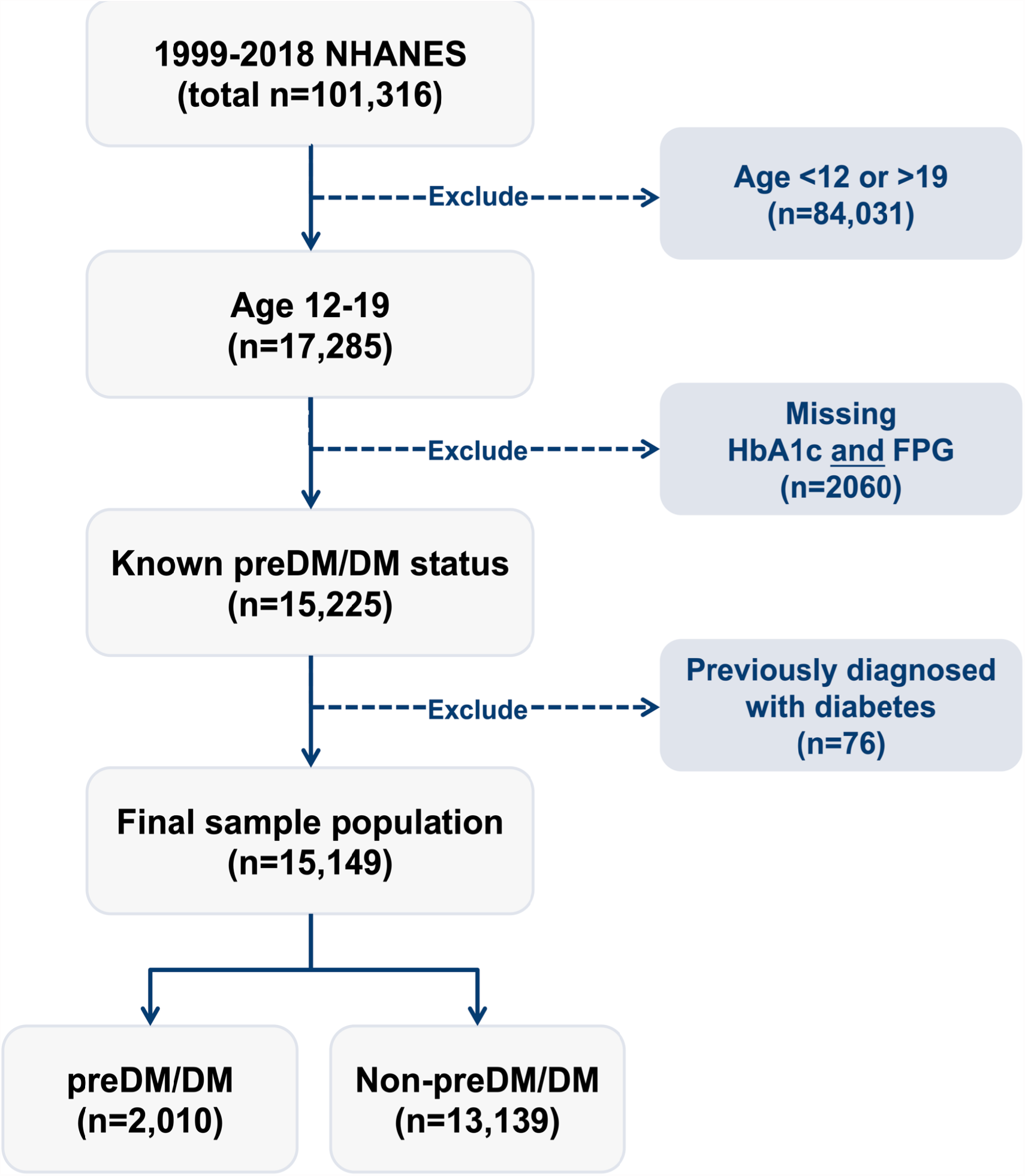
Flow chart showing the inclusion and exclusion criteria applied to 1999-2018 NHANES participants that yielded the study population included in our youth preDM/DM dataset. PreDM/DM status was defined by the current American Diabetes Association biomarker criteria, i.e., elevated levels of one of two preDM/DM biomarkers (fasting plasma glucose (FPG) ≥100 mg/dL or hemoglobin A1c (HbA1c) ≥ 5.7%). NHANES = National Health and Nutrition Examination Survey.

We extracted 95 epidemiological variables from NHANES, and organized them into four preDM/DM-related domains, namely sociodemographic, health status, diet, and other lifestyle behaviors **(Table S1). Table 1** shows the unweighted statistics of some key study population characteristics. Non-Hispanic black youth were disproportionately affected by preDM/DM as they comprised 33.6% of those with preDM/DM while representing only 27.4% of those without preDM/DM. Non-Hispanic white youth represented 21.4% of the preDM/DM youth as compared to 27.4% of the youth without preDM/DM. Hispanic youth showed similar proportions of those with and without preDM/DM at 35.4% and 36.9%, respectively. Youth categorized as Other represented 9.6% and 8.1% of those with and without preDM/DM, respectively. Approximately, half of the population were females, and they represented 34.4% of those with preDM/DM. Approximately 32.4% of the youth had a family income below poverty level, and 69.4% were from households receiving food stamps. The proportion of youth covered by private insurance was higher among those with than without preDM/DM (43.8% vs 37.7%). Overall, 21.5% of the youth were obese as defined by having a BMI at or above the 95th percentile based on age and gender, and the proportion was 33.3% among youth with preDM/DM. Youth with preDM/DM tended to have less fruit and vegetable intake and ate lower amounts of protein and total grains than those without. Youth with and without preDM/DM showed similar amounts of physical activity with 209 and 210 minutes per week, respectively **(Table 1)**.

### PreDM/DM in youth ONline Dashboard (POND)

To facilitate other researchers’ use of our youth preDM/DM dataset and make our methodology transparent and reproducible, we developed an interactive web portal named POND, https://rstudio-connect.hpc.mssm.edu/POND/. Users can navigate POND through its built-in functionalities. For example, users are able to explore the details of the 95 individual variables and their distributions by preDM/DM status, as well as examine the risk factors of youth preDM/DM identified from the case studies described below **(Fig. 3)**. POND also allows users to easily download the data to conduct their own analyses and explore other youth preDM/DM-related research questions. In addition, we make available all the code used to develop the dataset, our case studies, and POND itself.

**Fig. 3.**
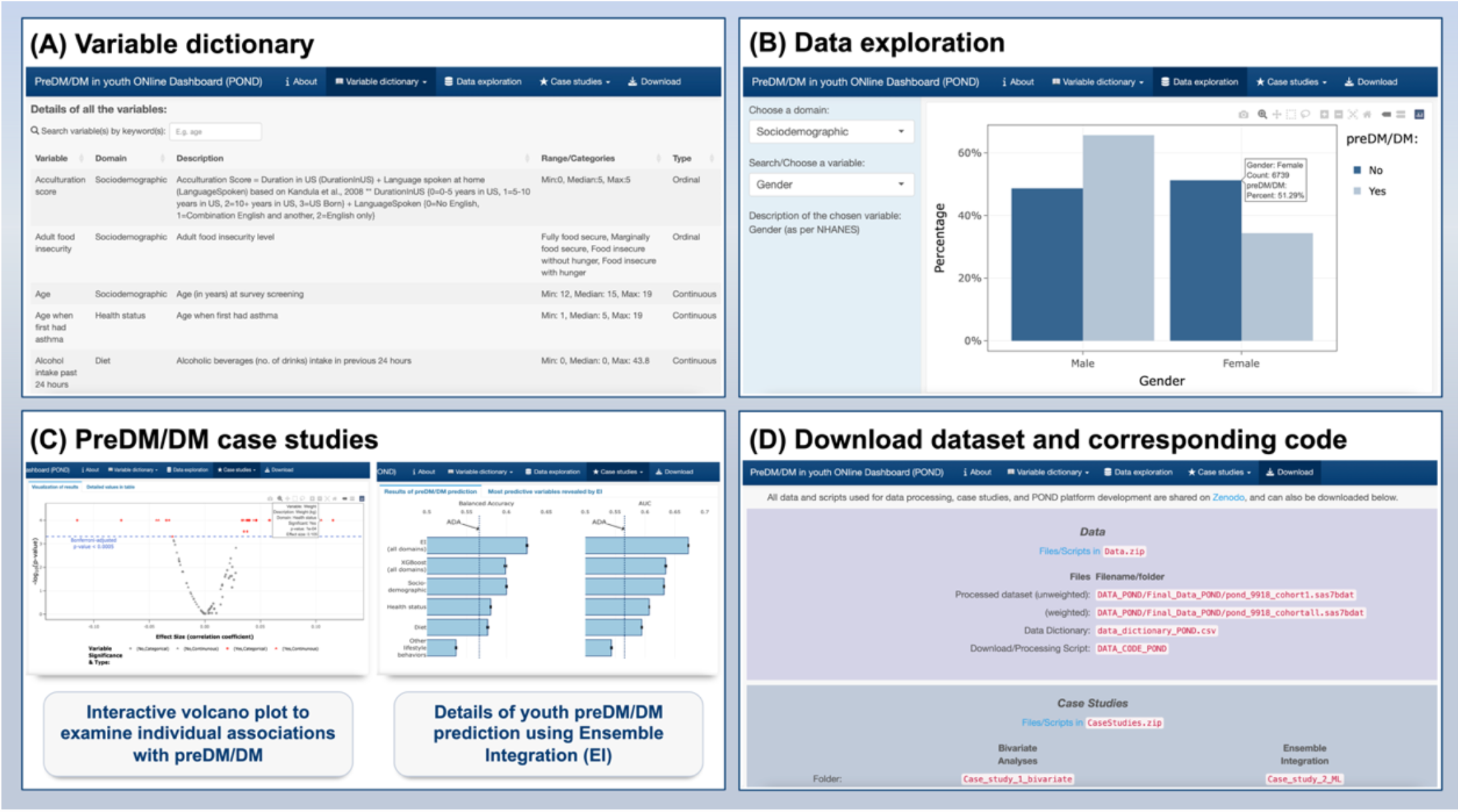
Screenshots of different functionalities available in POND (Prediabetes/diabetes in youth ONline Dashboard). **(A)** Detailed dictionary of the 95 variables included in our youth preDM/DM database organized by four domains, **(B)** Data exploration section showing the distribution of user selectable variables by preDM/DM status, **(C)** Case study section detailing the results of bivariate association analyses and the prediction of youth preDM/DM status from machine learning approaches and **(D)** Download section, where the dataset and the code used in the current study are publicly available to facilitate reproducibility and further exploration for interested users.

### Case studies using our dataset to better understand youth preDM/DM

We demonstrated the utility of the processed dataset for studying youth preDM/DM by two complementary types of data analyses. We first conducted exploratory bivariate analyses to investigate the statistical associations between individual variables and preDM/DM status using the Chi-squared and Wilcoxon rank-sum bivariate tests for categorical and continuous variables, respectively. In the second analysis, we examined the individual variable’s ability to predict preDM/DM status of youth using machine learning approaches. The results of these analyses are provided below.

#### Identifying individual variables associated with preDM/DM status

We found 27 variables to be significantly (p≤0.0005) associated with preDM/DM status, after Bonferroni adjustment for multiple testing **(Fig. 4, Table S1)**. These variables spanned all four domains, and included gender, race/ethnicity, use of food stamps, health insurance status, body mass index (BMI), total protein intake and screen time. Similar results were found when repeating these bivariate association tests after accounting for NHANES survey weights **(Table S1)**.

**Fig. 4.**
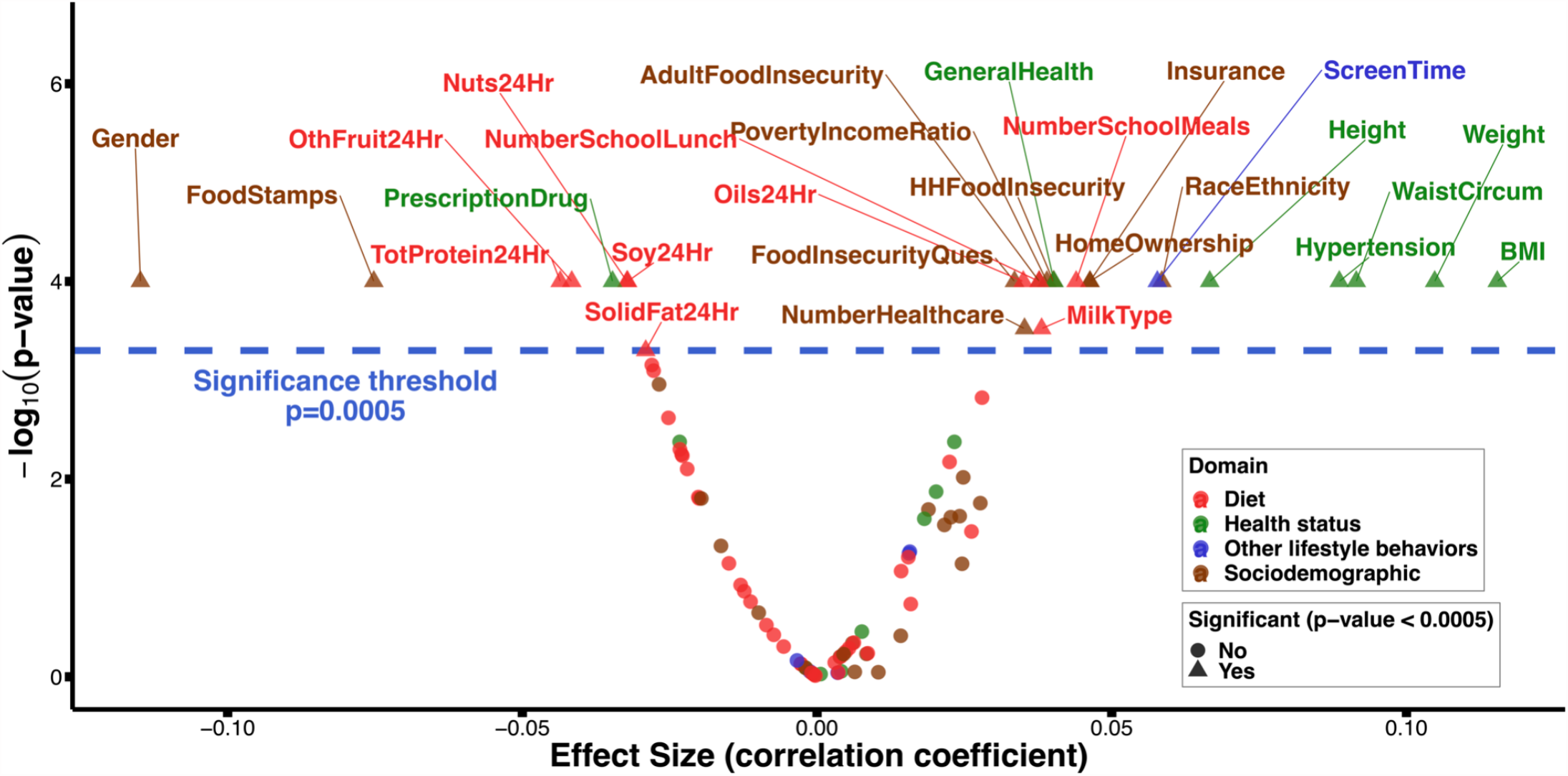
Individual variables associated with youth preDM/DM status based on bivariate analyses. This volcano plot shows the p-values and the effect sizes of the associations between the individual variables and youth preDM/DM status. Categorical and continuous variables were tested for association using Chi-square and Wilcoxon rank sum tests, respectively. Effect size was measured by Cramer’s V for categorical variables and Wilcoxon’s r-value (*73*) for continuous ones. After Bonferroni adjustment for multiple hypothesis testing, we found 27 variables to be significantly (p≤0.0005; blue dotted line) associated with youth preDM/DM status. These are named above the blue dotted line in this plot, and colored by the domain they belong to.

#### Predicting youth preDM/DM status

We used a machine learning framework, Ensemble Integration (EI) (*46*), which leverages the multi-domain nature of our dataset to predict youth preDM/DM status. We compared the predictive performance of EI with three alternative approaches (details in Materials and Methods): (i) A modified form of the American Academy of Pediatrics (AAP) and American Diabetes Association (ADA) screening guideline (*47*), (ii) single-domain derived EI predictors: sociodemographic, health status, diet and other lifestyle behaviors, and (iii) eXtreme Gradient Boosting (XGBoost) (*48*) using our full multi-domain dataset. The performance of EI and all the alternative approaches were assessed in terms of the commonly used Area Under the ROC Curve (AUC) (*49*) and Balanced Accuracy (BA, average of specificity and sensitivity) (*50*) measures **(Fig. 5)**. The performance of the machine learning-based prediction approaches, namely multi- and single-domain EI and XGBoost, were evaluated in a five-fold cross-validation setting repeated ten times (*51*). These performances were statistically compared using the Wilcoxon rank sum test, and the resultant p-values were corrected for multiple hypothesis testing using the Benjamini-Hochberg procedure to yield false discovery rates (FDRs) (*52*).

**Fig. 5.**
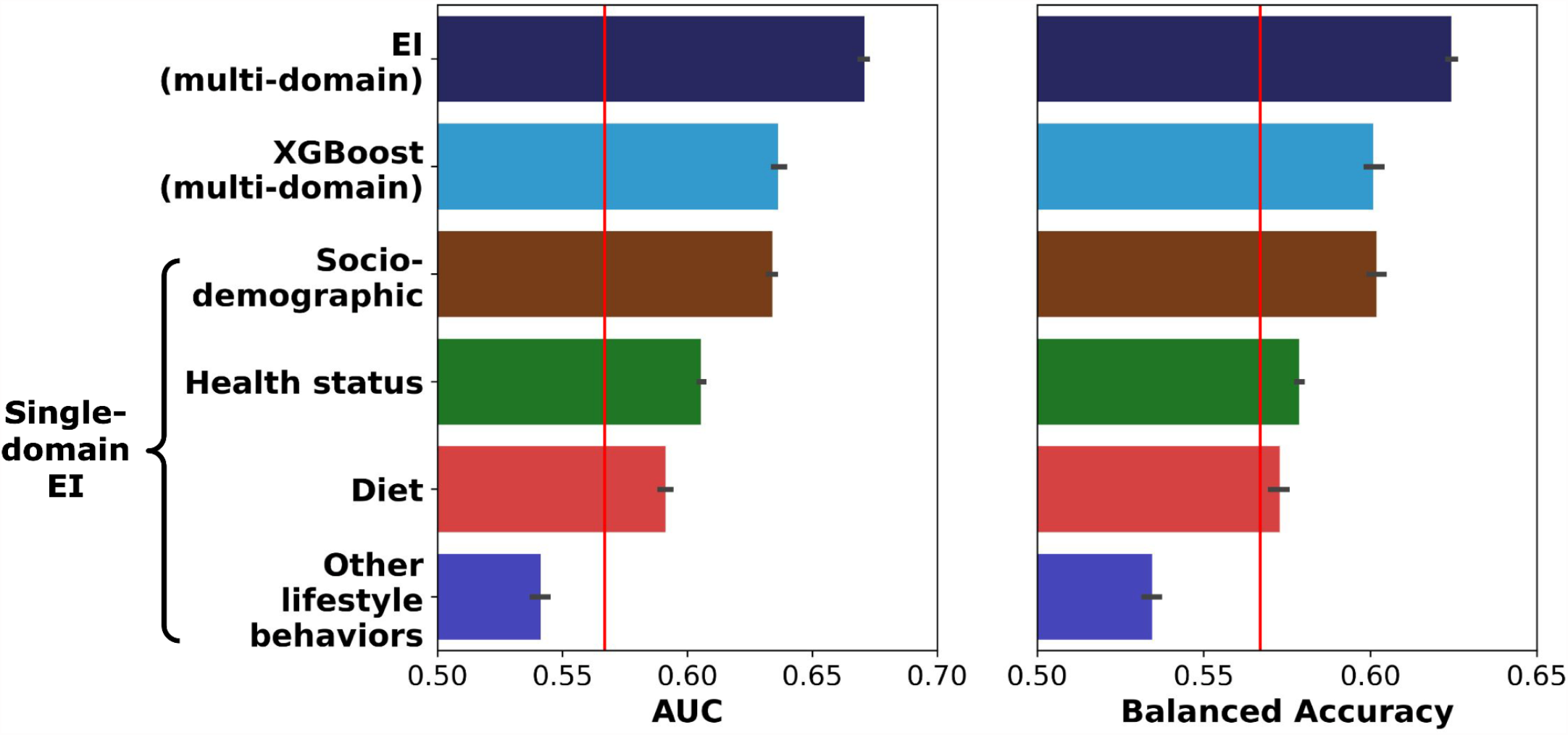
Comparison of the performance of multiple approaches for predicting youth preDM/DM status based on machine learning approaches. We compared the performance of the multi-domain Ensemble Integration (EI) approach with three alternative prediction approaches. The alternative approaches were: (i) a modified form of the American Academy of Pediatrics/American Diabetes Association screening guideline (vertical red line), (ii) single-domain EI-based prediction based on each of the four individual domains, and (iii) the commonly used eXtreme Gradient Boosting (XGBoost) algorithm applied to our whole dataset. Performance was measured in terms of the Area Under the ROC Curve (AUC) and Balanced Accuracy (average of sensitivity and specificity) measures. For each machine learning approach, the horizontal bar shows the average of the corresponding scores and the error bar indicates the corresponding standard error measured over ten rounds of five-fold cross-validation.

The best-performing multi-domain EI methodology, stacking (*53*) using Logistic Regression, predicted youth preDM/DM status (AUC=0.67, BA=0.62) more accurately than all the alternative approaches, namely XGBoost (AUC=0.64, BA=0.60, Wilcoxon rank-sum FDR=1.7×10^−4^ and 1.8×10^−4^, respectively), the ADA/AAP pediatric screening guidelines (AUC=0.57, BA=0.57; Wilcoxon rank-sum FDR=1.7×10^−4^ and 1.8×10^−4^, respectively), and EI applied to the four single domains (AUC=0.63-0.54, BA=0.60-0.53; FDR<1.7×10^−4^ and 1.8×10^−4^, respectively).

The multi-domain EI also identified 27 variables (the same as the number of significant variables from bivariate analyses) that contributed the most to predicting youth preDM/DM status. Among these variables, 17 overlapped with those identified from the bivariate statistical analyses (**Fig. 6**; Fisher’s p of overlap=7.06×10^−6^). These variables identified by both approaches included some established preDM/DM risk factors like BMI and high total cholesterol, as well as some less-recognized ones like screen time and taking prescription drugs (*2*).

**Fig. 6.**
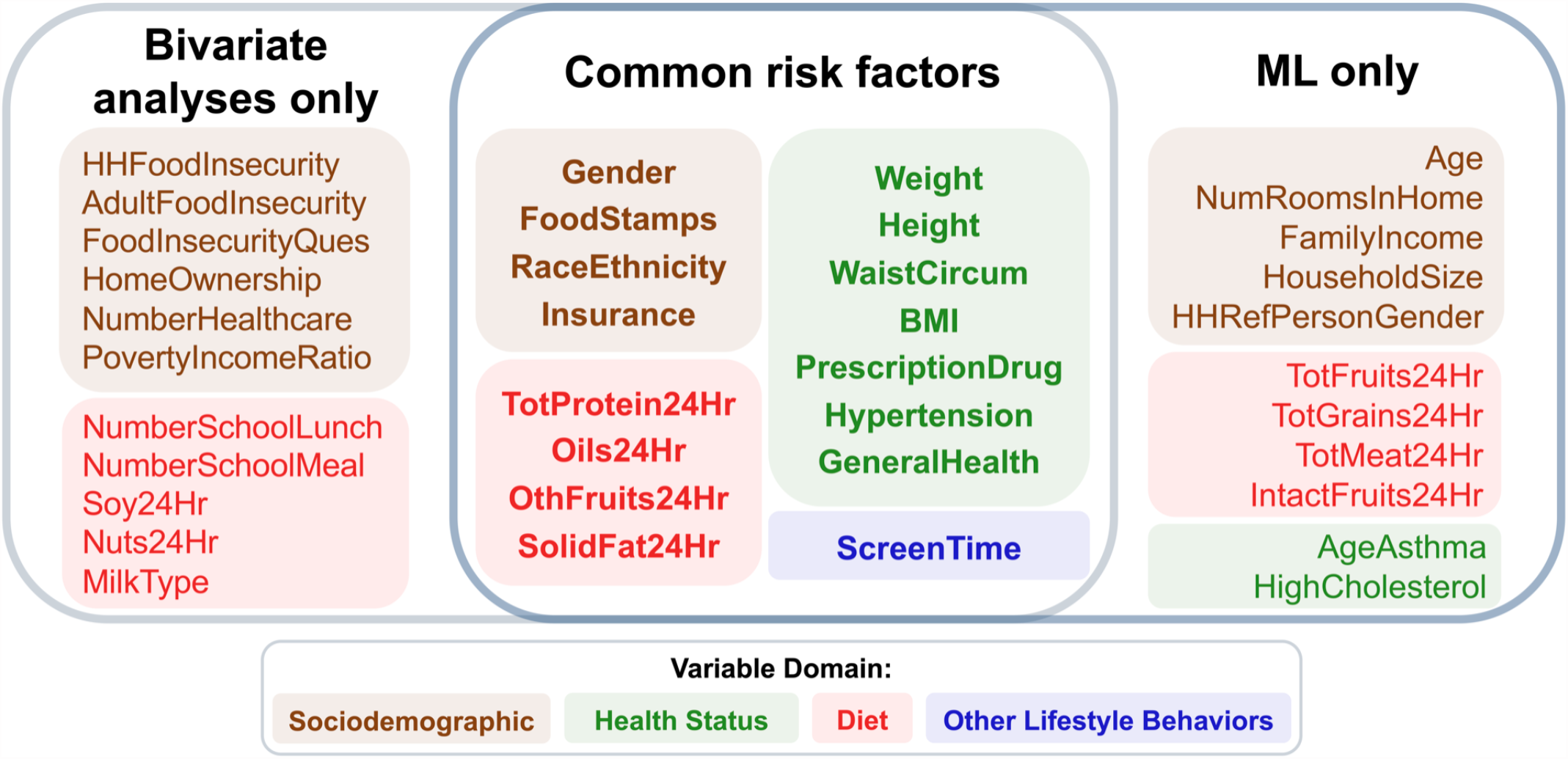
Variables associated with youth preDM/DM selected by bivariate analyses and the multi-domain EI approaches. Venn diagram summarizing the overlap between the 27 significant variables identified in the bivariate analyses and the 27 most predictive variables identified from the multidomain EI model. We found 16 variables overlapped between the two methods (Fisher’s p=7.06×10^−6^), and were drawn from all four domains (shown in different colors), indicating the multifactorial nature of youth preDM/DM.

## Discussion

Leveraging the rich information in NHANES spanning nearly 20 years, we built the most comprehensive epidemiological dataset for studying youth preDM/DM. We accomplished this by selecting and harmonizing variables relevant to youth preDM/DM from sociodemographic, health status, diet and other lifestyle behaviors domains. This youth preDM/DM dataset, as well as several functionalities to explore and analyze it, are publicly available in our user-friendly web portal, POND. We also conducted case studies using the dataset with both traditional statistical methods and machine learning approaches to demonstrate the potential of using this dataset to identify factors relevant to youth preDM/DM. The combination of the comprehensive public dataset and POND provide avenues for more informed investigations of youth preDM/DM.

The future impact of preDM/DM research, facilitated by comprehensive datasets like the one developed in this study, holds significant promise for advancing our understanding of the disease and its risk factors among youth. By enabling researchers to investigate multifactorial variables associated with preDM/DM, this dataset can contribute to several areas of research and have a broader impact on the scientific community. Firstly, the dataset’s comprehensive nature allows researchers to explore the collective impact of various risk factors across multiple health domains. By incorporating sociodemographic factors, health status indicators, diet, and lifestyle behaviors, researchers can gain a holistic understanding of the interplay between these factors and preDM/DM risk among youth. This knowledge can inform the development of targeted interventions and prevention strategies that address the specific needs of at-risk populations.

Furthermore, the dataset provides an opportunity to delve into less-studied variables and their interactions in relation to preDM/DM risk. Variables such as screen time, acculturation, or frequency of eating out, which are often overlooked in traditional research, can be examined to uncover their potential influence on preDM/DM risk among youth. This expands the scope of research and enhances our understanding of the multifaceted nature of the disease.

One of the major contributions of our work was POND, our publicly available web portal, which provided access to all materials related to our dataset and analyses, thus enabling transparency and reproducibility. Although several such portals are available in other biomedical areas, such as genomics (*54*–*56*), there is a general lack of such tools in epidemiology and public health. We hope that, in addition to facilitating studies into preDM/DM, POND illustrates the utility of such portals for these areas as well.

The results of the case studies we conducted are also consistent with existing literature, identifying known preDM/DM risk factors, such as gender (*11, 13*–*16*), race/ethnicity (*2, 7, 21, 23*), health measures (BMI, hypertension and cholesterol) (*2, 47*), income (*7, 8, 21*), insurance status (*7, 21*) and healthcare availability (*7, 21*), thus affirming the validity of the dataset. In addition, our analyses revealed some less studied variables, such as screen time, home ownership status, self-reported health status, soy and nut consumption, and frequency of school meal intake, that may influence youth preDM/DM risk. Further study of these variables may reveal new knowledge about preDM/DM among youth. More generally, such novel findings further demonstrate the utility of our dataset and data-driven methods for further discoveries about this complex disorder.

Although our work has several strengths and high potential utility for youth preDM/DM studies, it is not without limitations. First, as our dataset is derived from NHANES, we adopt limitations to the survey in our dataset. Since NHANES is a cross-sectional survey, the preDM/DM status and its related variables only provide consecutive snapshots of youth in the U.S. over time across the available survey cycles, and the associations identified are better suited for hypothesis generation purposes, which require in-depth investigation using prospective longitudinal and randomized trial designs. Additionally, we modified the APA/AAP guideline according to variable availability. Due to the high missingness of 45% in family history (DIQ170) and the complete missingness of maternal history (DIQ175S) from 1999-2010 in the raw NHANES data, we were unable to include family history of diabetes in the dataset. NHANES does not provide data regarding every condition associated with insulin resistance. Therefore, we used hypertension and high cholesterol as proxies for insulin resistance. On the other hand, as our main purpose is to use POND as a conduit between this comprehensive youth preDM/DM database and interested researchers, our method can be adopted to longitudinal data sets should they become available in the future. Second, for the prediction of preDM/DM status, EI’s performance was found to be significantly better than the alternative approaches, including a modified form of the suggested guideline (*44*). However, this performance assessment was only based on cross-validation, which is no substitute for validation on external datasets that is necessary for rigorous assessment. Finally, while our preliminary case study analyses identified a wide range of variables associated with youth prediabetes and diabetes, other known risk factors, such as current asthma status (*57*–*59*), added sugar consumption (*60*–*64*), sugary fruit and juice intake (*60*–*65*), and physical activity per week (*5, 71, 72*), were not identified. This limitation can be addressed by employing other data analysis methods beyond our bivariate testing and machine learning approaches, highlighting more potential use cases of our dataset.

Overall, the future impact of preDM/DM research facilitated by comprehensive datasets like ours extends beyond individual studies. It creates opportunities for interdisciplinary collaboration and reproducibility, strengthens evidence-based decision-making, and supports the development of targeted interventions for the prevention and management of preDM/DM among youth. By fostering a collaborative research environment, it enables researchers to build upon existing knowledge and push the boundaries of preDM/DM research, ultimately leading to improved health outcomes for at-risk populations.

## Materials and Methods

**Fig. 1** shows the overall study design and workflow. Below, we detail the components of the workflow.

### Data source and study population

We built the youth preDM/DM dataset based on NHANES data (*68*) spanning the years 1999 to 2018. Developed by the Centers for Disease Control and Prevention (CDC), NHANES is a serial cross-sectional survey that gathers comprehensive health-related information from nationally representative samples of the non-institutionalized population in the United States. The survey employs a multi-stage probability sampling method and collects data through questionnaires, physical examinations, and biomarker analysis. Each year, approximately 5,000 individuals are included in the survey, and the data are publicly released in 2-year cycles.

**Fig. 2** details the process used to define our study population. Briefly, of the total 101,316 participants in 1999-2018 NHANES, we excluded individuals who (i) were not within the 12–19-year age range, (ii) did not have either of the biomarkers used to define preDM/DM status, and (iii) answered, “Yes,” to “Have you ever been told by a doctor or health professional that you have diabetes?”. The final study population included 15,149 youth. Youth were considered at risk of preDM/DM if their Fasting Plasma Glucose (FPG) was at or greater than 100 mg/dL or their glycated hemoglobin (HbA1C) was at or greater than 5.7% according to the current ADA guidelines (*2*).

### Development of youth preDM/DM dataset

Based on the most recent ADA standard of care recommendations including factors related to preDM/DM risk and management (*2*), we selected 27 potentially relevant NHANES questionnaires and grouped them into four domains: sociodemographic, health status, diet, and other lifestyle behaviors. For example, under the health status domain, body mass index (BMI) was included as a potential risk factor for youth preDM/DM (*2*). Similarly, lifestyle and behavioral variables included factors, such as diet and physical activity, that have been shown to be critical for preDM/DM prevention in both observational studies and randomized clinical trials (*67, 69, 70*). Our sociodemographic domain included demographic variables and other social determinants of health (e.g., age, gender, poverty status, and food security). Except for commonly available clinical measurements, such as blood pressure and total cholesterol, we did not include laboratory data (e.g., triglycerides, transferrin, CRP, IL-6, WBC, etc.), since these measurements were not collected for all NHANES participants.

From the selected modules, we identified a list of 95 variables. The process of extracting these variables involved extensive examination of the questions that were asked, consultation of the literature, and discussions to reach consensus within the study team. The details of this process are provided in Section A of Supplemental Methods. We used SAS (version 9.4) and R (version 4.2.2; R Core Team, 2022) in R Studio (version 4.2.2; R Core Team, 2022) for data processing and dataset development. All the code developed and processed data are available in POND.

### Building the *pre*DM/DM in youth *ON*line *D*ashboard (POND)

We built POND to share our processed dataset and enable users to understand and explore the data on their own. The web portal was developed using R markdown and the flexdashboard package (*71*), and was published as a Shiny application (*72*). **Table S2** in Section B of Supplemental Methods provides details of all the R packages used to develop POND, and the related code is available on the portal’s download page.

### Case studies in using the dataset to better understand youth preDM/DM

To examine the utility of our dataset for studying youth preDM/DM, we conducted two complementary data analyses. We first conducted bivariate analyses to assess the statistical associations between each of the 95 variables and youth preDM/DM status. In the second analysis, we used machine learning methods to examine the ability to predict preDM/DM status of youth based on the 95 variables. The methodological details of these analyses are provided below.

#### Bivariate analyses to identify variables associated with preDM/DM status

We examined associations between individual variables and youth preDM/DM status using Chi-square and Wilcoxon rank sum tests for categorical and continuous variables, respectively. We applied Bonferroni correction for multiple hypothesis testing (n=95 tests) at an alpha level of 0.05 to determine the statistical significance of each association at the adjusted alpha level of 0.0005 (i.e., approximately 0.05/95). Finally, we used a volcano plot **(Fig. 4)** to visualize the results, where the y-axis is the log transformed p-value, and the x-axis is the effect size of the bivariate association. We used Cramer’s V and Wilcoxon R-values (*73*) as the effect size measures for categorical and continuous variables, respectively. To better compare with results from the machine learning approach, the main bivariate analyses did not account for NHANES survey design; thus, the results were only applicable to the study population included in the analytical sample, not generalizable to the entire U.S. youth population through survey weighting. For completeness, we provided the survey-weighted analyses in Section C of Supplemental Methods.

#### Prediction of preDM/DM status using machine learning algorithms

Several machine learning algorithms have been employed to predict adult preDM/DM status using NHANES data (*74*–*76*), and we have previously utilized these algorithms to predict preDM/DM status specifically among youth (*41*). However, to properly take into account the multi-domain nature of our dataset to build an effective and interpretable predictive model of youth preDM/DM, we leveraged our recent Ensemble Integration (EI) framework (*46*). EI incorporates both consensus and complementarity among the domains in our dataset by inferring local predictive models from the individual domains, and then integrating them into a global model using heterogeneous ensemble algorithms (*77*). EI also enables the identification of the most predictive variables in the final model, thus offering deeper insights into the outcome being predicted.

We used both the above capabilities of EI to build and interpret a predictive model of youth preDM/DM status based on our dataset. We also compared the predictive performance of the model with three alternative approaches: (i) a modified form of the AAP/ADA screening guideline (*47*), which is based on BMI, total cholesterol level, hypertension, and race/ethnicity, to assess the utility of data-driven screening for youth preDM/DM, (ii) EI applied to individual variable domains, namely sociodemographic, health status, diet and other lifestyle behaviors, to assess the value of multi-domain data for youth preDM/DM prediction, and (iii) eXtreme Gradient Boosting (XGBoost) (*48*) applied to our full dataset as a representative alternate machine learning algorithm that is considered the most effective for tabular data (*78*). More details of EI, the alternative approaches and the evaluation methodology, including cross-validation, model selection and comparison, are available in Section D of Supplemental Methods.

Finally, we used EI’s interpretation capabilities (*46*) to identify the variables in our dataset that were the most predictive of youth preDM/DM status. We selected the top 27 ranked predictors to compare to the 27 variables identified from the bivariate association analyses described above.

## Data Availability

The data and scripts used in this work were uploaded on Zenodo: https://zenodo.org/record/8206576. The web portal POND contains all scripts and data produced in the study and is hosted on https://rstudio-connect.hpc.mssm.edu/POND.

https://zenodo.org/record/8206576

## Acknowledgements

The study was enabled in part by computational resources provided by Scientific Computing and Data at the Icahn School of Medicine at Mount Sinai. The Ensemble Integration used in this work was implemented by Jamie J.R. Bennett.

## Author Contributions

Conceptualization: NV, BL, GP

Methodology: CM, YCL, NV, BL, GP

Software: CM, YCL, BL

Validation: CM, YCL, NV, BL, GP

Formal analysis: CM, YCL

Investigation: CM, YCL, NV, BL, GP

Resources: BL, GP

Data curation: CM, YCL

Writing – original draft: CM, YCL

Writing – review & editing: CM, YCL, NV, BL, GP

Visualization: CM, YCL

Supervision: NV, BL, GP

Project administration: NV, BL, GP

Funding acquisition: NV, BL, GP

## Data and materials availability

The data and scripts used in this work were uploaded on Zenodo: https://zenodo.org/record/8206576. The web portal POND is hosted on https://rstudio-connect.hpc.mssm.edu/POND.

## References

1. O. R. Temneanu, L. M. Trandafir, M. R. Purcarea, Type 2 diabetes mellitus in children and adolescents: a relatively new clinical problem within pediatric practice. J. Med. Life. 9, 235–239 (2016).

2. N. A. ElSayed, G. Aleppo, V. R. Aroda, R. R. Bannuru, F. M. Brown, D. Bruemmer, B. S. Collins, M. E. Hilliard, D. Isaacs, E. L. Johnson, S. Kahan, K. Khunti, J. Leon, S. K. Lyons, M. L. Perry, P. Prahalad, R. E. Pratley, J. J. Seley, R. C. Stanton, R. A. Gabbay, 2. Classification and Diagnosis of Diabetes: Standards of Care in Diabetes—2023. Diabetes Care. 46, S19–S40 (2023).

3. R. Weiss, S. Dufour, S. E. Taksali, W. V. Tamborlane, K. F. Petersen, R. C. Bonadonna, L. Boselli, G. Barbetta, K. Allen, F. Rife, M. Savoye, J. Dziura, R. Sherwin, G. I. Shulman, S. Caprio, Prediabetes in obese youth: a syndrome of impaired glucose tolerance, severe insulin resistance, and altered myocellular and abdominal fat partitioning. The Lancet. 362, 951–957 (2003).

4. Y. Zhang, A. O. Y. Luk, E. Chow, G. T. C. Ko, M. H. M. Chan, M. Ng, A. P. S. Kong, R. C. W. Ma, R. Ozaki, W. Y. So, C. C. Chow, J. C. N. Chan, High risk of conversion to diabetes in first-degree relatives of individuals with young-onset type 2 diabetes: a 12-year follow-up analysis. Diabet. Med. J. Br. Diabet. Assoc. 34, 1701–1709 (2017).

5. P. Zhuang, X. Liu, Y. Li, X. Wan, Y. Wu, F. Wu, Y. Zhang, J. Jiao, Effect of Diet Quality and Genetic Predisposition on Hemoglobin A1c and Type 2 Diabetes Risk: Gene-Diet Interaction Analysis of 357,419 Individuals. Diabetes Care. 44, 2470–2479 (2021).

6. J. A. Pivovarov, C. E. Taplin, M. C. Riddell, Current perspectives on physical activity and exercise for youth with diabetes: Perspectives on exercise. Pediatr. Diabetes. 16, 242–255 (2015).

7. F. Hill-Briggs, N. E. Adler, S. A. Berkowitz, M. H. Chin, T. L. Gary-Webb, A. Navas-Acien, P. L. Thornton, D. Haire-Joshu, Social Determinants of Health and Diabetes: A Scientific Review. Diabetes Care. 44, 258–279 (2021).

8. R. J. Walker, B. L. Smalls, J. A. Campbell, J. L. Strom Williams, L. E. Egede, Impact of social determinants of health on outcomes for type 2 diabetes: a systematic review. Endocrine. 47, 29–48 (2014).

9. N. Bansal, Prediabetes diagnosis and treatment: A review. World J. Diabetes. 6, 296 (2015).

10. T. Tönnies, R. Brinks, S. Isom, D. Dabelea, J. Divers, E. J. Mayer-Davis, J. M. Lawrence, C. Pihoker, L. Dolan, A. D. Liese, S. H. Saydah, R. B. D. Jr., A. Hoyer, G. Imperatore, “Projections of type 1 and type 2 diabetes burden in the US population aged <20 years through 2060: The SEARCH for Diabetes in Youth Study” (other, 2022),, doi:10.2337/figshare.21514014.

11. J. M. Lawrence, J. Divers, S. Isom, S. Saydah, G. Imperatore, C. Pihoker, S. M. Marcovina, E. J. Mayer-Davis, R. F. Hamman, L. Dolan, D. Dabelea, D. J. Pettitt, A. D. Liese, SEARCH for Diabetes in Youth Study Group, Trends in Prevalence of Type 1 and Type 2 Diabetes in Children and Adolescents in the US, 2001-2017. JAMA. 326, 717 (2021).

12. E. T. Jensen, D. Dabelea, Type 2 Diabetes in Youth: New Lessons from the SEARCH Study. Curr. Diab. Rep. 18, 36 (2018).

13. D. Dabelea, E. J. Mayer-Davis, S. Saydah, G. Imperatore, B. Linder, J. Divers, R. Bell, A. Badaru, J. W. Talton, T. Crume, A. D. Liese, A. T. Merchant, J. M. Lawrence, K. Reynolds, L. Dolan, L. L. Liu, R. F. Hamman, Prevalence of Type 1 and Type 2 Diabetes Among Children and Adolescents From 2001 to 2009. JAMA. 311, 1778 (2014).

14. N. Lascar, J. Brown, H. Pattison, A. H. Barnett, C. J. Bailey, S. Bellary, Type 2 diabetes in adolescents and young adults. Lancet Diabetes Endocrinol. 6, 69–80 (2018).

15. L. J. Andes, Y. J. Cheng, D. B. Rolka, E. W. Gregg, G. Imperatore, Prevalence of Prediabetes Among Adolescents and Young Adults in the United States, 2005-2016. JAMA Pediatr. 174, e194498 (2020).

16. A. Menke, S. Casagrande, C. C. Cowie, Prevalence of Diabetes in Adolescents Aged 12 to 19 Years in the United States, 2005-2014. JAMA. 316, 344–345 (2016).

17. M. A. B. Khan, M. J. Hashim, J. K. King, R. D. Govender, H. Mustafa, J. Al Kaabi, Epidemiology of Type 2 Diabetes – Global Burden of Disease and Forecasted Trends. J. Epidemiol. Glob. Health. 10, 107–111 (2020).

18. X. Lin, Y. Xu, X. Pan, J. Xu, Y. Ding, X. Sun, X. Song, Y. Ren, P.-F. Shan, Global, regional, and national burden and trend of diabetes in 195 countries and territories: an analysis from 1990 to 2025. Sci. Rep. 10, 14790 (2020).

19. G. Imperatore, J. P. Boyle, T. J. Thompson, D. Case, D. Dabelea, R. F. Hamman, J. M. Lawrence, A. D. Liese, L. L. Liu, E. J. Mayer-Davis, B. L. Rodriguez, D. Standiford, SEARCH for Diabetes in Youth Study Group, Projections of type 1 and type 2 diabetes burden in the U.S. population aged <20 years through 2050: dynamic modeling of incidence, mortality, and population growth. Diabetes Care. 35, 2515–2520 (2012).

20. J. Liu, Y. Li, D. Zhang, S. S. Yi, J. Liu, Trends in Prediabetes Among Youths in the US From 1999 Through 2018. JAMA Pediatr. 176, 608–611 (2022).

21. A. M. Butler, Social Determinants of Health and Racial/Ethnic Disparities in Type 2 Diabetes in Youth. Curr. Diab. Rep. 17, 60 (2017).

22. W. H. Herman, Y. Ma, G. Uwaifo, S. Haffner, S. E. Kahn, E. S. Horton, J. M. Lachin, M. G. Montez, T. Brenneman, E. Barrett-Connor, for the Diabetes Prevention Program Research Group, Differences in A1C by Race and Ethnicity Among Patients With Impaired Glucose Tolerance in the Diabetes Prevention Program. Diabetes Care. 30, 2453–2457 (2007).

23. A. R. Kahkoska, C. M. Shay, J. Crandell, D. Dabelea, G. Imperatore, J. M. Lawrence, A. D. Liese, C. Pihoker, B. A. Reboussin, S. Agarwal, J. A. Tooze, L. E. Wagenknecht, V. W. Zhong, E. J. Mayer-Davis, Association of Race and Ethnicity With Glycemic Control and Hemoglobin A1c Levels in Youth With Type 1 Diabetes. JAMA Netw. Open. 1, e181851 (2018).

24. A. M. Lee, C. R. Fermin, S. L. Filipp, M. J. Gurka, M. D. DeBoer, Examining trends in prediabetes and its relationship with the metabolic syndrome in US adolescents, 1999-2014. Acta Diabetol. 54, 373–381 (2017).

25. R. Weiss, S. E. Taksali, W. V. Tamborlane, T. S. Burgert, M. Savoye, S. Caprio, Predictors of changes in glucose tolerance status in obese youth. Diabetes Care. 28, 902–909 (2005).

26. K. J. Nadeau, B. J. Anderson, E. G. Berg, J. L. Chiang, H. Chou, K. C. Copeland, T. S. Hannon, T. T.-K. Huang, J. L. Lynch, J. Powell, E. Sellers, W. V. Tamborlane, P. Zeitler, Youth-Onset Type 2 Diabetes Consensus Report: Current Status, Challenges, and Priorities. Diabetes Care. 39, 1635–1642 (2016).

27. A. B. Dart, P. J. Martens, C. Rigatto, M. D. Brownell, H. J. Dean, E. A. Sellers, Earlier Onset of Complications in Youth With Type 2 Diabetes. Diabetes Care. 37, 436–443 (2014).

28. American Diabetes Association, Economic Costs of Diabetes in the U.S. in 2017. Diabetes Care. 41, 917–928 (2018).

29. A. S. Al-Goblan, M. A. Al-Alfi, M. Z. Khan, Mechanism linking diabetes mellitus and obesity. Diabetes Metab. Syndr. Obes. Targets Ther. 7, 587–591 (2014).

30. J. C. N. Chan, L.-L. Lim, N. J. Wareham, J. E. Shaw, T. J. Orchard, P. Zhang, E. S. H. Lau, B. Eliasson, A. P. S. Kong, M. Ezzati, C. A. Aguilar-Salinas, M. McGill, N. S. Levitt, G. Ning, W.-Y. So, J. Adams, P. Bracco, N. G. Forouhi, G. A. Gregory, J. Guo, X. Hua, E. L. Klatman, D. J. Magliano, B.-P. Ng, D. Ogilvie, J. Panter, M. Pavkov, H. Shao, N. Unwin, M. White, C. Wou, R. C. W. Ma, M. I. Schmidt, A. Ramachandran, Y. Seino, P. H. Bennett, B. Oldenburg, J. J. Gagliardino, A. O. Y. Luk, P. M. Clarke, G. D. Ogle, M. J. Davies, R. R. Holman, E. W. Gregg, The Lancet Commission on diabetes: using data to transform diabetes care and patient lives. The Lancet. 396, 2019–2082 (2020).

31. International Diabetes Federation, IDF Diabetes Atlas, 10th Edition, (available at https://diabetesatlas.org/).

32. U.S. Chronic Disease Indicators: Diabetes | Chronic Disease and Health Promotion Data & Indicators, (available at https://chronicdata.cdc.gov/Chronic-Disease-Indicators/U-S-Chronic-Disease-Indicators-Diabetes/f8ti-h92k).

33. NCD Risk Factor Collaboration, (available at https://ncdrisc.org/index.html).

34. International Diabetes Federation, Diabetes Atlas., World Bank Open Data - Diabetes prevalence. World Bank Open Data, (available at https://data.worldbank.org/indicator/SH.STA.DIAB.ZS).

35. B. Zhou, et al., Worldwide trends in diabetes since 1980: a pooled analysis of 751 population-based studies with 4·4 million participants. The Lancet. 387, 1513–1530 (2016).

36. UCI Machine Learning Repository: Diabetes 130-US hospitals for years 1999-2008 Data Set, (available at https://archive.ics.uci.edu/ml/datasets/Diabetes+130-US+hospitals+for+years+1999-2008).

37. Type 2 Diabetes Knowledge Portal, (available at https://t2d.hugeamp.org/).

38. A. Rashid, Diabetes Dataset. 1 (2020), doi:10.17632/wj9rwkp9c2.1.

39. Diabetes Dataset 2019, (available at https://www.kaggle.com/datasets/tigganeha4/diabetes-dataset-2019).

40. Diabetes Health Indicators Dataset, (available at https://www.kaggle.com/datasets/alexteboul/diabetes-health-indicators-dataset).

41. N. Vangeepuram, B. Liu, P. Chiu, L. Wang, G. Pandey, Predicting youth diabetes risk using NHANES data and machine learning. Sci. Rep. 11, 11212 (2021).

42. S. Nagarajan, A. Khokhar, D. S. Holmes, S. Chandwani, Family Consumer Behaviors, Adolescent Prediabetes and Diabetes in the National Health and Nutrition Examination Survey (2007-2010). J. Am. Coll. Nutr. 36, 520–527 (2017).

43. A. S. Wallace, D. Wang, J.-I. Shin, E. Selvin, Screening and Diagnosis of Prediabetes and Diabetes in US Children and Adolescents. Pediatrics. 146, e20200265 (2020).

44. P. Chu, A. Patel, V. Helgeson, A. B. Goldschmidt, M. K. Ray, M. E. Vajravelu, Perception and Awareness of Diabetes Risk and Reported Risk-Reducing Behaviors in Adolescents. JAMA Netw. Open. 6, e2311466 (2023).

45. C. J. Patel, N. Pho, M. McDuffie, J. Easton-Marks, C. Kothari, I. S. Kohane, P. Avillach, A database of human exposomes and phenomes from the US National Health and Nutrition Examination Survey. Sci. Data. 3, 160096 (2016).

46. Y. C. Li, L. Wang, J. N. Law, T. M. Murali, G. Pandey, Integrating multimodal data through interpretable heterogeneous ensembles. Bioinforma. Adv. 2, vbac065 (2022).

47. S. Arslanian, F. Bacha, M. Grey, M. D. Marcus, N. H. White, P. Zeitler, Evaluation and Management of Youth-Onset Type 2 Diabetes: A Position Statement by the American Diabetes Association. Diabetes Care. 41, 2648–2668 (2018).

48. T. Chen, C. Guestrin, “XGBoost: A Scalable Tree Boosting System” in Proceedings of the 22nd ACM SIGKDD International Conference on Knowledge Discovery and Data Mining (Association for Computing Machinery, New York, NY, USA, 2016; https://dl.acm.org/doi/10.1145/2939672.2939785), KDD ‘16, pp. 785–794.

49. N. Altman, M. Krzywinski, Graphical assessment of tests and classifiers. Nat. Methods. 18, 840–842 (2021).

50. K. H. Brodersen, C. S. Ong, K. E. Stephan, J. M. Buhmann, “The Balanced Accuracy and Its Posterior Distribution” in 2010 20th International Conference on Pattern Recognition (2010), pp. 3121–3124.

51. S. Arlot, A. Celisse, A survey of cross-validation procedures for model selection. Stat. Surv. 4, 40–79 (2010).

52. Y. Benjamini, Y. Hochberg, Controlling the False Discovery Rate: A Practical and Powerful Approach to Multiple Testing. J. R. Stat. Soc. Ser. B Methodol. 57, 289–300 (1995).

53. M. P. Sesmero, A. I. Ledezma, A. Sanchis, Generating ensembles of heterogeneous classifiers using Stacked Generalization. WIREs Data Min. Knowl. Discov. 5, 21–34 (2015).

54. P. L. Whetzel, N. F. Noy, N. H. Shah, P. R. Alexander, C. Nyulas, T. Tudorache, M. A. Musen, BioPortal: enhanced functionality via new Web services from the National Center for Biomedical Ontology to access and use ontologies in software applications. Nucleic Acids Res. 39, W541–545 (2011).

55. S. Bhattacharya, S. Andorf, L. Gomes, P. Dunn, H. Schaefer, J. Pontius, P. Berger, V. Desborough, T. Smith, J. Campbell, E. Thomson, R. Monteiro, P. Guimaraes, B. Walters, J. Wiser, A. J. Butte, ImmPort: disseminating data to the public for the future of immunology. Immunol. Res. 58, 234–239 (2014).

56. J. Zhang, J. Baran, A. Cros, J. M. Guberman, S. Haider, J. Hsu, Y. Liang, E. Rivkin, J. Wang, B. Whitty, M. Wong-Erasmus, L. Yao, A. Kasprzyk, International Cancer Genome Consortium Data Portal—a one-stop shop for cancer genomics data. Database. 2011, bar026 (2011).

57. L. Rayner, A. McGovern, B. Creagh-Brown, C. Woodmansey, S. de Lusignan, Type 2 Diabetes and Asthma: Systematic Review of the Bidirectional Relationship. Curr. Diabetes Rev. 15, 118–126 (2019).

58. M. H. Black, A. Anderson, R. A. Bell, D. Dabelea, C. Pihoker, S. Saydah, M. Seid, D. A. Standiford, B. Waitzfelder, S. M. Marcovina, J. M. Lawrence, Prevalence of Asthma and Its Association With Glycemic Control Among Youth With Diabetes. Pediatrics. 128, e839–e847 (2011).

59. T. D. Wu, Diabetes, insulin resistance, and asthma: a review of potential links. Curr. Opin. Pulm. Med. 27, 29–36 (2021).

60. D. C. Greenwood, D. E. Threapleton, C. E. L. Evans, C. L. Cleghorn, C. Nykjaer, C. Woodhead, V. J. Burley, Association between sugar-sweetened and artificially sweetened soft drinks and type 2 diabetes: systematic review and dose-response meta-analysis of prospective studies. Br. J. Nutr. 112, 725–734 (2014).

61. L. R. Vartanian, M. B. Schwartz, K. D. Brownell, Effects of Soft Drink Consumption on Nutrition and Health: A Systematic Review and Meta-Analysis. Am. J. Public Health. 97, 667–675 (2007).

62. V. S. Malik, B. M. Popkin, G. A. Bray, J.-P. Després, W. C. Willett, F. B. Hu, Sugar-sweetened beverages and risk of metabolic syndrome and type 2 diabetes: a meta-analysis. Diabetes Care. 33, 2477–2483 (2010).

63. D. P. DiMeglio, R. D. Mattes, Liquid versus solid carbohydrate: effects on food intake and body weight. Int. J. Obes. Relat. Metab. Disord. J. Int. Assoc. Study Obes. 24, 794–800 (2000).

64. B. M. Popkin, Patterns of beverage use across the lifecycle. Physiol. Behav. 100, 4–9 (2010).

65. I. Muraki, F. Imamura, J. E. Manson, F. B. Hu, W. C. Willett, R. M. van Dam, Q. Sun, Fruit consumption and risk of type 2 diabetes: results from three prospective longitudinal cohort studies. BMJ. 347, f5001 (2013).

66. S. R. Colberg, R. J. Sigal, J. E. Yardley, M. C. Riddell, D. W. Dunstan, P. C. Dempsey, E. S. Horton, K. Castorino, D. F. Tate, Physical Activity/Exercise and Diabetes: A Position Statement of the American Diabetes Association. Diabetes Care. 39, 2065–2079 (2016).

67. A. Sampath Kumar, A. G. Maiya, B. A. Shastry, K. Vaishali, N. Ravishankar, A. Hazari, S. Gundmi, R. Jadhav, Exercise and insulin resistance in type 2 diabetes mellitus: A systematic review and meta-analysis. Ann. Phys. Rehabil. Med. 62, 98–103 (2019).

68. G. Zipf, M. Chiappa, K. S. Porter, Y. Ostchega, B. G. Lewis, J. Dostal, National health and nutrition examination survey: plan and operations, 1999-2010. Vital Health Stat. Ser 1 Programs Collect. Proced., 1–37 (2013).

69. K. Karstoft, C. S. Christensen, B. K. Pedersen, T. P. J. Solomon, The acute effects of interval-Vs continuous-walking exercise on glycemic control in subjects with type 2 diabetes: a crossover, controlled study. J. Clin. Endocrinol. Metab. 99, 3334–3342 (2014).

70. K. Karstoft, K. Winding, S. H. Knudsen, J. S. Nielsen, C. Thomsen, B. K. Pedersen, T. P. J. Solomon, The Effects of Free-Living Interval-Walking Training on Glycemic Control, Body Composition, and Physical Fitness in Type 2 Diabetic Patients. Diabetes Care. 36, 228–236 (2013).

71. R Markdown Format for Flexible Dashboards, (available at https://pkgs.rstudio.com/flexdashboard/).

72. Shiny - Welcome to Shiny, (available at https://shiny.posit.co/r/getstarted/shiny-basics/lesson1/index.html).

73. M. Tomczak, E. Tomczak, The need to report effect size estimates revisited. An overview of some recommended measures of effect size. Trends Sport Sci. 1, 19–25 (2014).

74. W. H. Herman, P. J. Smith, T. J. Thompson, M. M. Engelgau, R. E. Aubert, A new and simple questionnaire to identify people at increased risk for undiagnosed diabetes. Diabetes Care. 18, 382–387 (1995).

75. H. Bang, A. M. Edwards, A. S. Bomback, C. M. Ballantyne, D. Brillon, M. A. Callahan, S. M. Teutsch, A. I. Mushlin, L. M. Kern, Development and validation of a patient self-assessment score for diabetes risk. Ann. Intern. Med. 151, 775–783 (2009).

76. E. Poltavskiy, D. J. Kim, H. Bang, Comparison of screening scores for diabetes and prediabetes. Diabetes Res. Clin. Pract. 118, 146–153 (2016).

77. S. Whalen, O. P. Pandey, G. Pandey, Predicting protein function and other biomedical characteristics with heterogeneous ensembles. Methods San Diego Calif. 93, 92–102 (2016).

78. R. Shwartz-Ziv, A. Armon, Tabular data: Deep learning is not all you need. Inf. Fusion. 81, 84–90 (2022).

