## Supplementary Materials for "Facilitating youth diabetes studies with the most comprehensive epidemiological dataset available through a public web portal"

Catherine McDonough *et al.*

**The PDF file includes:**
Tables S1 to S4
Figs. S1 to S3
Appendices A to D
References 47, 50, 52, 68, 79 to 84

### Supplemental Results

**Table S1. Details of the 95 epidemiological variables included in the youth preDM/DM dataset.** Variables that were found to be statistically significantly (p≤0.0005) associated with youth preDM/DM status in both weighted and unweighted bivariate analyses are shown in bold. This information can also be accessed through the Variable dictionary and Case studies sections of POND.

| Variable | Domain | Type | N | Unweighted bivariate p-value | Weighted bivariate p-value |
|---|---|---|---|---|---|
| Acculturation score | Sociodemographic | Ordinal | 14,947 | 0.8992§ | 0.0100§ |
| **Adult food insecurity** | Sociodemographic | Ordinal | 14,760 | **0.0001** | **<0.0001** |
| Age | Sociodemographic | Continuous | 15,149 | 0.0157 | 0.0542 |
| Attend kindergarten through high school | Sociodemographic | Binary | 15,142 | 0.9188 | 0.9777 |
| **Authorized or received food stamps** | Sociodemographic | Binary | 11,294 | **0.0001** | **<0.0001** |
| Chemical use in lawn/garden | Sociodemographic | Binary | 14,776 | 0.0474 | 0.0928 |
| Child food insecurity | Sociodemographic | Ordinal | 13,234 | 0.0175 | **0.0004** |
| Country of birth | Sociodemographic | Binary | 15,143 | 0.6287 | 0.3333 |
| Education level | Sociodemographic | Ordinal | 15,142 | 0.0096 | 0.0734 |
| Family income | Sociodemographic | Ordinal | 14,177 | 0.0719 | 0.1780 |
| **Food insecurity questions** | Sociodemographic | Binary | 14,758 | **0.0001** | **<0.0001** |
| **Gender** | Sociodemographic | Categorical | 15,149 | **0.0001** | **<0.0001** |
| **Health insurance** | Sociodemographic | Ordinal | 14,876 | **0.0001** | **<0.0001** |
| **Home ownership** | Sociodemographic | Categorical | 14,944 | **0.0001** | **0.0001** |
| **Household food insecurity** | Sociodemographic | Ordinal | 14,760 | **0.0001** | **<0.0001** |
| Household reference person age | Sociodemographic | Ordinal | 15,149 | 0.3847 | 0.2981 |
| Household reference person education level | Sociodemographic | Ordinal | 14,485 | 0.0243 | 0.0020 |
| Household reference person gender | Sociodemographic | Categorical | 15,145 | 0.0203 | 0.2995 |
| Household WIC received | Sociodemographic | Binary | 14,390 | 0.5901 | 0.3487 |
| Languages spoken at home | Sociodemographic | Ordinal | 15,021 | 0.5868 | **0.0004** |
| Nativity acculturation score | Sociodemographic | Ordinal | 15,072 | 0.8924 | 0.7035 |
| Number of people in household | Sociodemographic | Ordinal | 15,149 | 0.0291 | 0.0127 |
| Number of rooms in home | Sociodemographic | Continuous | 14,919 | 0.0011 | 0.0119 |
| **Number of times received healthcare last year** | Sociodemographic | Ordinal | 15,117 | **0.0003** | **0.0001** |
| Overnight hospitalization last year | Sociodemographic | Binary | 15,145 | 0.2247 | 0.1001 |
| **Race and Ethnicity** | Sociodemographic | Categorical | 15,149 | **0.0001** | **<0.0001** |
| **Ratio of family income to poverty** | Sociodemographic | Ordinal | 13,956 | **0.0001** | **0.0002** |
| Routine healthcare visit | Sociodemographic | Binary | 15,145 | 0.8148 | 0.4066 |
| Type of healthcare visit the most | Sociodemographic | Categorical | 12,723 | 0.0237 | 0.0011 |
| US citizenship | Sociodemographic | Binary | 15,116 | 0.4570 | 0.3569 |
| Age when first had asthma* | Health status | Continuous | 15,092 | 0.0042 | 0.0300 |
| Anemia Treatment | Health status | Binary | 15,143 | 0.8060 | 0.7067 |
| **BMI** | Health status | Ordinal | 14,980 | **0.0001** | **<0.0001** |
| Current asthma | Health status | Binary | 15,096 | 0.0252 | 0.5732 |
| **General Health Condition** | Health status | Ordinal | 15,146 | **0.0001** | **<0.0001** |
| Health changes | Health status | Categorical | 15,146 | 0.8803 | 0.7353 |
| High cholesterol level | Health status | Ordinal | 14,912 | 0.0134 | 0.0942 |
| **Hypertension** | Health status | Ordinal | 14,643 | **0.0001** | **<0.0001** |
| Lifetime asthma | Health status | Binary | 15,135 | 0.0042 | 0.0548 |
| Mental health visit | Health status | Binary | 15,140 | 0.9388 | 0.6696 |
| Received Blood Transfusions | Health status | Binary | 15,122 | 0.3491 | 0.6780 |
| **Standing Height** | Health status | Continuous | 15,010 | **0.0001** | **<0.0001** |
| **Taken prescription drugs last month** | Health status | Binary | 15,142 | **0.0001** | **0.0002** |
| **Waist Circumference** | Health status | Continuous | 14,794 | **0.0001** | **<0.0001** |
| **Weight** | Health status | Continuous | 14,985 | **0.0001** | **<0.0001** |
| Alcohol intake past 24 hours† | Diet | Continuous | 14,549 | 0.0056 | 0.0006 |
| Cheese intake past 24 hours† | Diet | Continuous | 14,549 | 0.4946 | 0.6670 |

| | | | | | |
|---|---|---|---|---|---|
| Citrus and melons and berries intake past 24 hours† | Diet | Continuous | 14,549 | 0.1364 | 0.5268 |
| Cured meat intake past 24 hours† | Diet | Continuous | 14,549 | 0.0617 | 0.4717 |
| Dark green vegetables intake past 24 hours† | Diet | Continuous | 14,549 | 0.9117 | 0.9131 |
| Eggs intake past 24 hours† | Diet | Continuous | 14,549 | 0.0067 | 0.0349 |
| Foods added sugars intake past 24 hours† | Diet | Continuous | 14,549 | 0.1179 | 0.6739 |
| High n3 fatty acids seafood intake past 24 hours† | Diet | Continuous | 14,549 | 0.7452 | 0.7172 |
| Legume intake past 24 hours† | Diet | Continuous | 14,549 | 0.0157 | 0.7901 |
| Low n3 fatty acids seafood intake past 24 hours† | Diet | Continuous | 14,549 | 0.3762 | 0.5440 |
| Meals out of home per week† | Diet | Continuous | 14,890 | 0.6209 | 0.3536 |
| Milk intake past 24 hours† | Diet | Continuous | 14,549 | 0.0050 | **0.0005** |
| **Number of meals from School‡** | Diet | Continuous | 14,264 | **0.0001** | 0.4383 |
| Milk intake past 30 days | Diet | Ordinal | 15,141 | 0.0339 | 0.0140 |
| **Nuts intake past 24 hours†** | Diet | Continuous | 14,549 | **0.0001** | 0.1595 |
| **Oils intake past 24 hours†** | Diet | Continuous | 14,549 | **0.0001** | 0.0017 |
| Organ meat intake past 24 hours† | Diet | Continuous | 14,549 | 0.9539 | 0.5493 |
| **Other fruits intake past 24 hours†** | Diet | Continuous | 14,549 | **0.0001** | 0.0052 |
| Other red vegetables intake past 24 hours† | Diet | Continuous | 14,549 | 0.9723 | 0.8675 |
| Other starchy vegetables intake past 24 hours† | Diet | Continuous | 14,549 | 0.5580 | 0.2494 |
| Other vegetables intake past 24 hours† | Diet | Continuous | 14,549 | 0.0079 | 0.0295 |
| Poultry intake past 24 hours† | Diet | Continuous | 14,549 | 0.0853 | 0.1643 |
| Red meat intake past 24 hours† | Diet | Continuous | 14,549 | 0.0008 | 0.0322 |
| Refined or non whole grains intake past 24 hours | Diet | Continuous | 14,549 | 0.1737 | 0.6316 |
| School breakfast per week‡ | Diet | Continuous | 12,835 | 0.0015 | **0.0001** |
| **School lunch per week‡** | Diet | Continuous | 14,467 | **0.0001** | 0.0092 |
| School with complete breakfast | Diet | Categorical | 14,899 | 0.5778 | 0.9642 |
| School with complete breakfast or lunch per day | Diet | Binary | 15,033 | 0.9064 | 0.9093 |
| School with complete lunch | Diet | Categorical | 13,396 | 0.1837 | 0.6588 |
| **Solid fat intake past 24 hours** | Diet | Continuous | 14,549 | **0.0005** | 0.0151 |
| **Soy products intake past 24 hours** | Diet | Continuous | 14,549 | **0.0001** | 0.6877 |
| Tomatoes and oranges intake past 24 hours† | Diet | Continuous | 14,549 | 0.0058 | 0.1235 |
| Tomatoes intake past 24 hours† | Diet | Continuous | 14,549 | 0.0024 | 0.0185 |
| Total dairy intake past 24 hours† | Diet | Continuous | 14,549 | 0.0152 | 0.0472 |
| Total fruits intake past 24 hours† | Diet | Continuous | 14,549 | 0.0007 | 0.0517 |
| Total meat intake past 24 hours† | Diet | Continuous | 14,549 | 0.7170 | 0.5028 |
| **Total protein intake past 24 hours†** | Diet | Continuous | 14,549 | **0.0001** | **0.0001** |
| Total starchy vegetables intake past 24 hours† | Diet | Continuous | 14,549 | 0.5130 | 0.9164 |
| Total vegetables intake past 24 hours† | Diet | Continuous | 14,549 | 0.0712 | 0.0877 |
| Total whole and refined grains intake past 24 hours | Diet | Continuous | 14,549 | 0.3001 | 0.5318 |
| **Type of milk drink** | Diet | Ordinal | 14,781 | **0.0003** | 0.0007 |
| White potatoes intake past 24 hours† | Diet | Continuous | 14,549 | 0.4515 | 0.9757 |
| Whole grains intake past 24 hours† | Diet | Continuous | 14,549 | 0.9450 | 0.7343 |
| Yogurt intake past 24 hours | Diet | Continuous | 14,549 | 0.7382 | 0.1720 |
| Exposed to secondhand smoke at home | Other lifestyle behaviors | Binary | 15,041 | 0.0565 | 0.0026 |
| Number of smokers in home† | Other lifestyle behaviors | Continuous | 14,963 | 0.0542 | 0.0046 |
| Physical activity per week† | Other lifestyle behaviors | Continuous | 14,792 | 0.8836 | 0.3612 |
| Physical limitation | Other lifestyle behaviors | Ordinal | 15,109 | 0.9096 | 0.2633 |
| Recent tobacco use | Other lifestyle behaviors | Binary | 14,248 | 0.6831 | 0.5971 |
| **Screen time per day** | Other lifestyle behaviors | Continuous | 14,781 | **0.0001** | **<0.0001** |

*For those without asthma, AgeAsthma is coded as 100. Prior to running survey-weighted regression, these were calculated as missing.

†In preparation for survey-weighted analysis, this variable was determined to have a skewness ≤-2 or ≥2, and therefore was unfit for linear regression. It was log-transformed prior to performing regression.

‡In survey-weighted analysis, this variable showed a bimodal distribution and was converted to binary indicator whose significance was determined with logistic regression.

§p-value from Chi-square tests for categorical variables and Wilcoxon rank sum tests for continuous variables, with Bonferroni adjustment for multiple hypothesis testing (adjusted alpha level= 0.05/95=0.0005). Survey weighted analyses took into account NHANES survey design elements (primary sampling unit, strata, and survey weight).

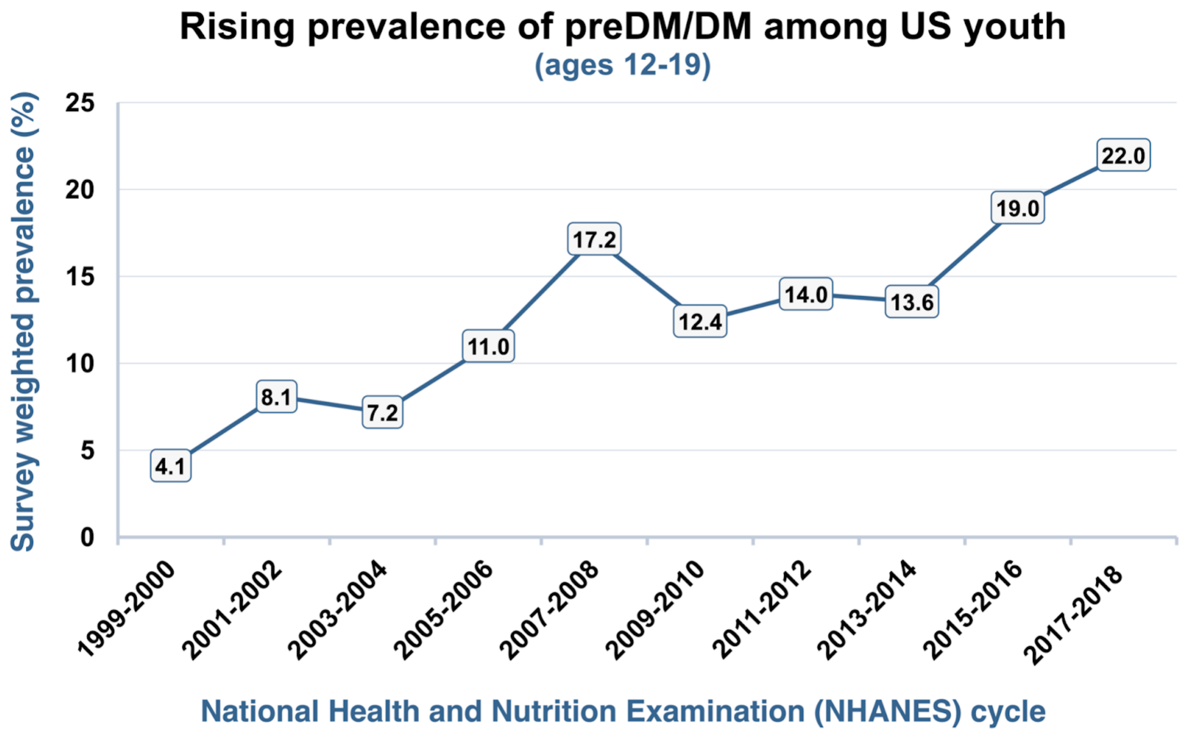


**Figure S1. Rising prevalence of preDM/DM among US youth based on 1999-2018 NHANES data.**

**Appendix A. Generating our youth prediabetes/diabetes (preDM/DM) dataset.**

We used a four-step process to generate the youth preDM/DM dataset from NHANES **(Fig. S2)**, as detailed below:

**Step 0: Download raw NHANES data**

We downloaded raw data from the NHANES website (https://wwwn.cdc.gov/nchs/nhanes/Default.aspx), which were organized by individual questionnaires and 2-year cycles. We organized these raw data into four domains relevant to youth preDM/DM: sociodemographic, health status, diet, and other lifestyle behaviors.

**Step 1: Select youth population**

We selected 15,149 youth study population based on three criteria **(Fig. 2 in the main text)**, as described in the main text.

**Step 2: Data pre-processing**

We pre-processed the 21 individual questionnaires from the raw NHANES data. This process consisted of cleaning existing variables, combining comparable variables, recategorizing values of comparable variables, combining hierarchical variables, and composite variable building, all of which are detailed below:

1. *Cleaning existing variables*. For variables that remained consistent across all cycles, we coded responses such as "Refused" or "Don't Know" as missing.
2. *Combining comparable variables*. To ensure consistency in variables with wording or name changes, a new variable was created by merging comparable values from different survey cycles.
3. *Recategorizing values of comparable variables*. When the wording of a question changed across cycles and/or the corresponding possible responses to the same question differed, we recoded the variables to keep consistent response levels. For example, in 2011, NHANES introduced the category of "Non-Hispanic Asian" in the race/ethnicity variable, which was not present in earlier cycles. Here, Non-Hispanic Asian individuals were recategorized as "Other" to align with the values in earlier survey cycles.
4. *Utilizing other variables to fill in missing hierarchical variables*. Hierarchical variables were identified when the answer to a higher-level question could be used to fill in missing information for a more specific question. For instance, if someone responded "No" to the question about ever having asthma, the subsequent question regarding their current asthma status would be skipped. Thus, for the current asthma variable, individuals who never had asthma were coded as "does not currently have asthma."
5. *Composite variable building*. Some variables were built from many variables that may not have matched perfectly. For example, for food stamps, we assumed that "being authorized in the last year" (1999-2006) and "receiving food stamps ever" (2007-2018) were treated as equivalent. Therefore, individuals who answered "Yes" to either were coded as yes for authorized or received food stamps.
6. *Third party resources*. We used validated third party resources to recode certain variables as recommended standard practice. For example, we applied the SAS Program for CDC Growth Charts to obtain BMI percentiles *(1)*, the Childhood Blood Pressure Macro to estimate blood pressure percentiles (*2*), and the United States Department of Agriculture (USDA) Food Patterns Equivalents Database (FPED) and MyPyramid Patterns Equivalents Database (MPED) to calculate daily food group intake (*3*).

**Step 3: Compile cleaned data into the analytical dataset**

Finally, we generated a master analytical database with all cleaned 95 variables organized by their corresponding domains (sociodemographic, health status, diet, and other lifestyle behaviors) for the 15,149 youth study population. In addition, a companion database for analyses for incorporating NHANES survey design elements were also generated, which included the 15,149 youth and participants who did not meet our selection criteria. The final youth preDMDM datasets serve as the foundation for further analysis and exploration of youth preDM/DM trends and patterns.

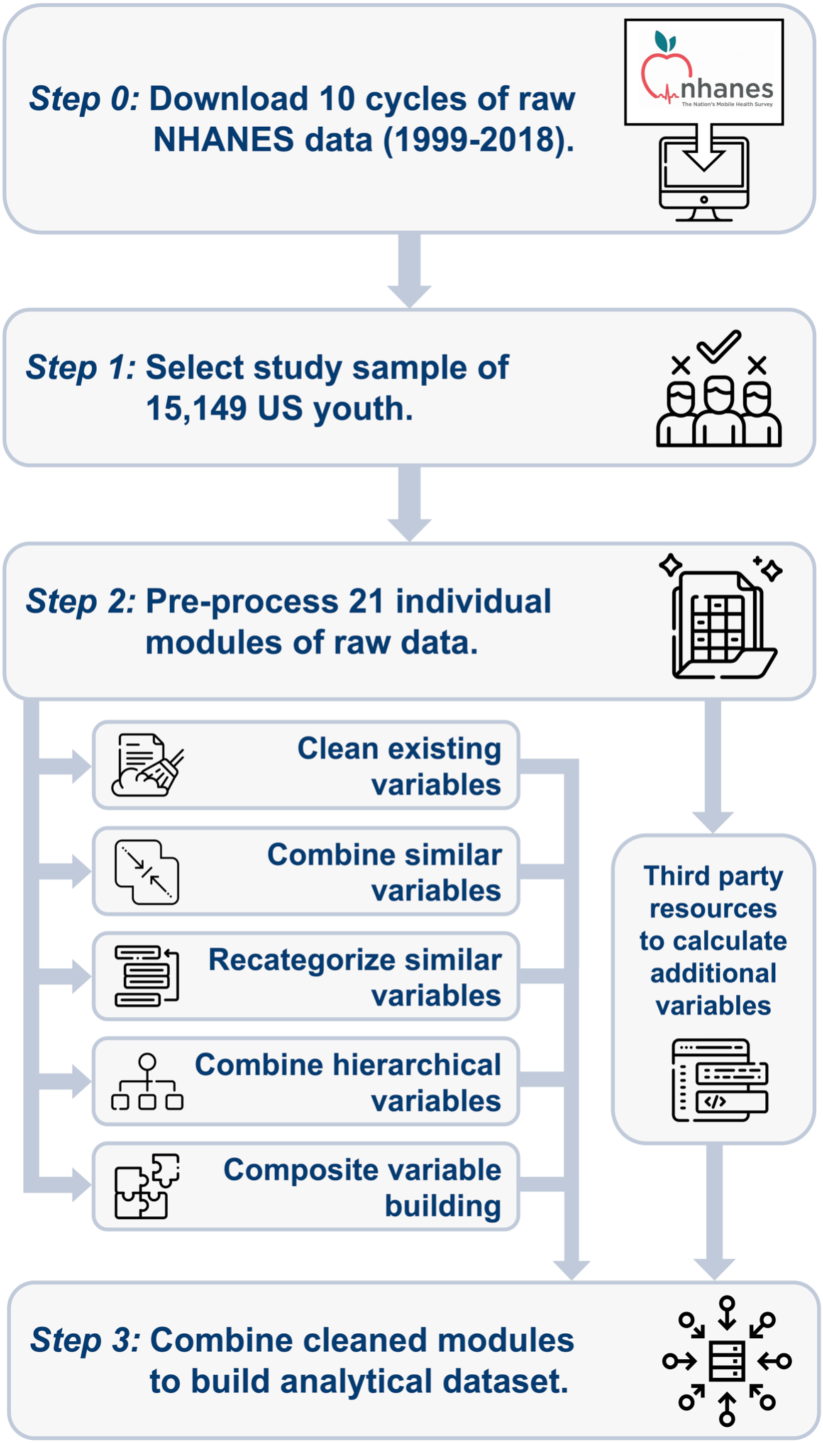


**Figure S2. Details of processing and standardizing raw NHANES variables to develop our youth preDM/DM dataset.** (Some images in this figure were obtained from the open-source collection at https://www.flaticon.com and were made by Freepik.)

### Appendix B. Implementation details of POND, our user-friendly data portal

The complete code implementing POND can be obtained from the portal's download page. The entire portal is coded in 'index.Rmd', which is a Rmarkdown file composed mainly of R, markdown, and a few HTML and Javascript codes. We used flexdashboard and shiny package to make the Rmarkdown file into the shiny web application in order to facilitate interactive and accessible online user experience. Several other packages are used to enrich the interactive components of POND, such as DT to show the table of variables with a search functionality, tidyverse and dplyr to load and process the youth preDM/DM dataset as a dataframe, and ggplot2 and plotly to draw dynamic and interactive plots. The complete list of packages used and their purpose are listed in **Table S2**.

**Table S2. R packages and their versions used to build POND.**

| R package | Version | Purpose |
|---|---|---|
| flexdashboard | 0.6.0 | Build the user interface of web portal |
| ggplot2 | 3.4.0 | Plot histogram, box whisker plot in Data Exploration & volcaCorrelates of preDM/DM status |
| reshape2 | 1.4.4 | Transform dataframe shapes (wide to long) |
| scales | 1.2.1 | Change the axis ticks of plots |
| tidyverse | 1.3.2 | Data analytic package |
| plotly | 4.10.1 | Make plot from ggplot2 into interactive, and make barplot of machine learning performance |
| dplyr | 1.0.10 | Process dataframes |
| shiny | 1.6.0 | Make R markdown document to be interactive |
| jsonlite | 1.7.2 | Read JSON files |
| DT | 0.27 | Show the variable dictionary and table of p-values |
| patchwork | 1.1.2 | Combine subfigures into a larger figure |

### Appendix C. Survey-weighted bivariate analyses

We conducted similar comparisons of individual variables by youth preDM/DM status using survey-weighted procedures. NHANES utilizes a multi-stage probability sampling scheme to generate a sample that is nationally representative of the non-institutionalized civilian U.S. population (i.e., the target population) (*4*). In order to produce population-level inference with results that are generalizable, the analyses need to incorporate the survey elements (e.g., primary sampling unit (PSU), strata, and survey sampling weight). Otherwise, the results are only applicable to the specific individuals included in the analytical sample. To that end, we conducted survey-weighted bivariate Rao-Scott Chi-Square tests using proc surveyfreq for categorical variables and survey-weighted linear regression models using proc surveyreg for continuous variables using SAS (version 9.4). NHANES examination weights (WTMEC2YR) were applied in all procedures. We applied Bonferroni correction for multiple hypothesis testing (n=95 tests), and a p-value less or equal to than 0.0005 (i.e., 0.05/95) was considered statistically significant.

To better fit the survey-weighted linear regression model, the following variables were natural-log-transformed to better approximate normality: PAminWk, MealsOut, GWhole, VTot, VDrkGr, VRedOrTot, VRedOr, VTomato, VStarchTot, VStarchPot, VStarchOth, VOther, Legume, FTot, FCitMlB, FOther, DTot, DMilk, DYogurt, DCheese, PTot, Pmps, PMeat, PFrank, POrgan, PPoult, PFishLow, PFishHigh, PEgg, PSoy, PNut, Oils, AddSugars, Alcohol, and HHSmkNum. Additionally, three variables (NwkBkfast, NwkLunch, and NwkMeals) exhibited bimodal distributions. The code used for survey-weighted bivariate analyses is available for download from POND.

### <u>Appendix D. Details of the approaches used to predict youth preDM/DM status</u>

#### Ensemble Integration (EI)

EI used in this work adopted the Python implementation of the original framework (*5*) **(Fig. S3)**. Specifically, we applied ten different base prediction algorithms **(Table S3)** to each of the domains in our dataset to develop local models. Next, EI models were built over the local models using heterogeneous ensemble methods, specifically mean, iterative ensemble selection method (*6*, *7*) and the ten stacking classifiers listed in **Table S4**.

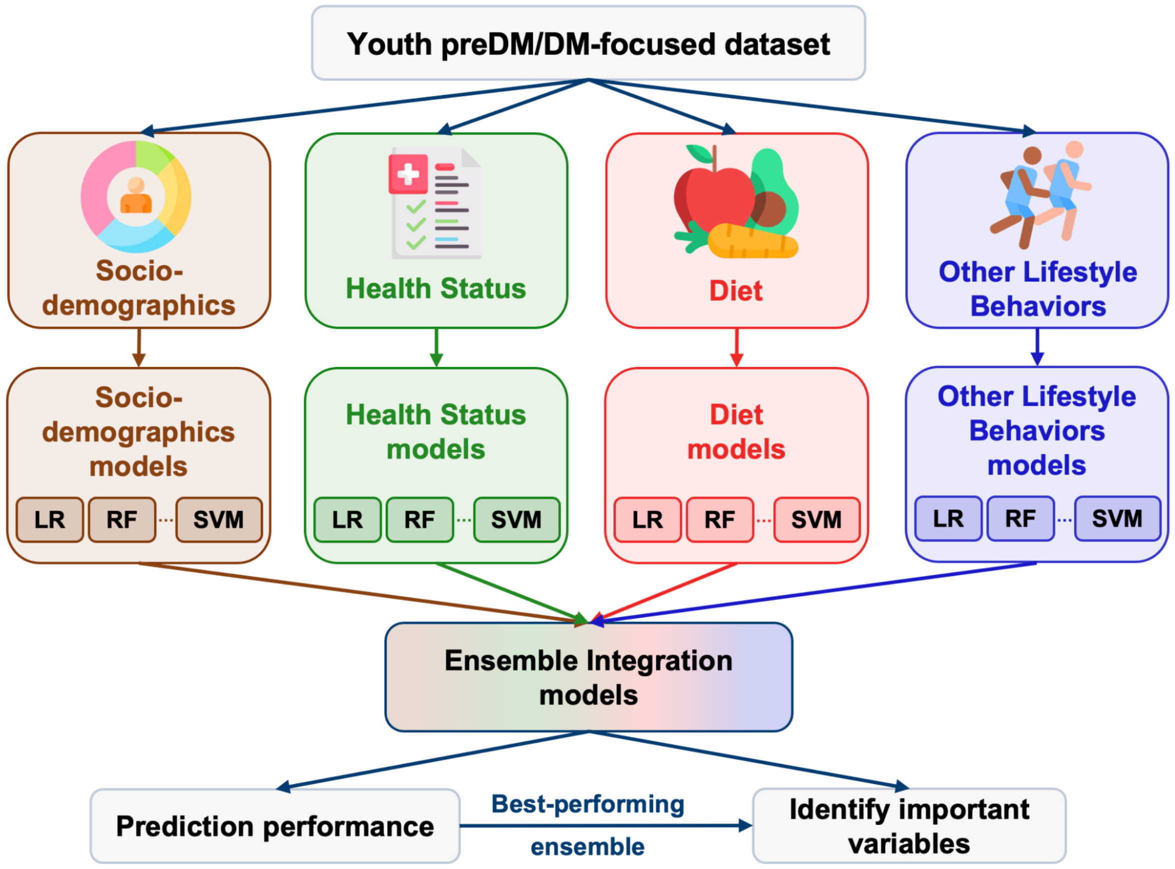


**Figure S3. Workflow of Ensemble Integration (EI)-based prediction of youth preDM/DM from our multi-domain dataset.**

First, ten standard binary classification algorithms listed in **Table S3**, such as Logistic Regression (LR), Random Forest (RF) and Support Vector Machine (SVM), were used to derive local domain-specific models. Heterogeneous ensemble methods were then applied to these domain-specific models to generate the EI models. These models generated preDM/DM prediction scores for every participant in our dataset, which were evaluated to assess the model prediction performance. Finally, we used the best-performing EI model to report performance results and identify the variables important for predicting youth preDM/DM status. (Some images in this figure were obtained from the open-source collection at https://www.flaticon.com and were made by Freepik.)

**Table S3. Base predictors used in EI to derive local domain-specific models.**

| Algorithm | Python class |
|---|---|
| Naive Bayes | sklearn.naive_bayes.GaussianNB() |
| Logistic Regression | make_pipeline(StandardScaler(), sklearn.linear_model.LogisticRegression()) |
| Support Vector Machine | make_pipeline(Normalizer(), sklearn.svm.SVC(kernel='poly', degree=1, probability=True)) |
| Perceptron | sklearn.linear_modelPerceptron() |
| AdaBoost | sklearn.ensemble.AdaBoostClassifier(n_estimators=10) |
| Decision Tree | sklearn.tree.DecisionTreeClassifier() |
| Gradient Boosting | sklearn.ensemble.GradientBoostingClassifier() |
| Random Forest | sklearn.ensemble.RandomForestClassifier() |
| Decision Tree from XGBoost | xgboost.XGBClassifier(n_estimators=1) |
| K Nearest Neighbors | sklearn.neighbors.KNeighborsClassifier(n_neighbors=1) |

**Table S4. Stacking classifiers used to generate final EI models.**

| Algorithm | Python class |
|---|---|
| Random Forest | sklearn.ensemble.RandomForestClassifier() |
| Support Vector Machine | sklearn.svm.SVC(kernel='linear', probability=True, max_iter=1e7) |
| Naive Bayes | sklearn.naive_bayes.GaussianNB() |
| Logistic Regression | sklearn.linear_model.LogisticRegression() |
| AdaBoost | sklearn.ensemble.AdaBoostClassifier(n_estimators=10) |
| Decision Tree | sklearn.tree.DecisionTreeClassifier() |
| Gradient Boosting | sklearn.ensemble.GradientBoostingClassifier() |
| Random Forest | sklearn.ensemble.RandomForestClassifier() |
| XGBoost | xgboost.XGBClassifier() |
| K Nearest Neighbors | sklearn.neighbors.KNeighborsClassifier() |

### Data Imputation

Several of the base classification algorithms in EI, such as SVM, cannot operate with missing values. Thus, we imputed the missing values according to variable types. For binary, categorical and ordinal variables, we imputed with the most frequent non-missing value (mode). For continuous variables, we imputed with the median of the non-missing values. The missing percentage range was 0-25.4% for binary, categorical and ordinal variables, and was 0-15.3% for continuous variables.

### Modified AAP/ADA guideline

The original American Academy of Pediatrics/American Diabetes Association guideline (*8*) used five variables to specify youth at risk for preDM/DM, namely being overweight (BMI ≥ 85th percentile for age and sex, weight for height ≥ 85th percentile, or weight ≥ 120% of ideal for height), combined with one of the following: (i) signs of insulin resistance or conditions associated with insulin resistance **(Table S5)**, (ii) race/ethnicity of Native American, African American, Latino, Asian American, or Pacific Islander, (iii) family history of diabetes of type 2 diabetes in first- or second-degree relative, or (iv) maternal history of diabetes or gestational diabetes during the mother's pregnancy with the child. Due to the high missingness of 45% in family history (DIQ170) and the missingness of maternal history (DIQ175S) from 1999-2010 in the raw NHANES data, we were only able to use four variables available in our youth preDM/DM dataset, which were BMI, total cholesterol level, hypertension, race/ethnicity **(Table S5)**. We further restricted the prediction to the sub-sample where none of these four variables were missing (14,292 participants). The modified guideline for determining whether an individual was at risk for preDM/DM was as follows: an individual was considered to be at risk for preDM/DM if they were overweight/obese (BMI ≥ 85% percentile) and satisfied at least one of the following conditions: total cholesterol level ≥ 170 mg/dL, hypertension, whose criterion is detailed in **Table S5**, or belonging to a non-White race/ethnicity.

**Table S5. Modified pediatric clinical screening guideline currently used for defining preDM/DM status and equivalent NHANES variables.**

| ADA/AAP preDM/DM risk for children (at-risk if overweight plus one or more additional risk factors) | NHANES variables used (at-risk if overweight plus one or more additional risk factors) |
|---|---|
| Overweight (BMI >85th percentile for age and sex, weight for height >85th percentile, or weight >120% of ideal for height) **A*** | BMI ≥ 85th percentile |
| *Additional risk factors:* | |
| Race/Ethnicity (Native American, African American, Latinx, Asian American, Pacific Islander) **A*** | Non-white race/ethnicity (non-Hispanic Black, Hispanic, other) |
| Signs of insulin resistance or conditions associated with insulin resistance (hypertension, dyslipidemia, acanthosis nigricans, polycystic ovary syndrome, or small-for-gestational-age birth weight) **B*** | Hypertension: Blood pressure ≥ 90th percentile or 120/80 mm Hg for children ≥ 13 years old; Dyslipidemia: total cholesterol ≥ 170 mg/dL |

*Evidence grades, with grade A and B representing higher and moderate quality evidence, respectively. In the current study, these were not factored into preDM/DM definition.

**Details of the evaluation of the prediction methods**

The performances of the machine learning (ML) methods were evaluated over ten runs of five-fold cross-validation on the whole dataset. Both the AAP/ADA screening guideline, as well as the ML methods tested produced scores for positive (+) and negative (-) preDM/DM status. The threshold of scores to determine the final preDM/DM status was obtained by maximizing Balanced Accuracy (BA), the average of sensitivity and specificity (*9*). For multi-domain EI on the all-domains combined full dataset, as well as the individual-domain EIs and XGBoost methods, only the implementations with the highest average BA over the ten cross-validation runs were considered in the comparison. We used the Wilcoxon rank sum test to assess the statistical significance of the comparison of the predictive performances of all ML methods tested, as well as the modified screening guideline. The resultant p-values were corrected for multiple hypothesis testing using the Benjamini-Hochberg method (*10*), which yielded the false discovery rates (FDRs) reported in this paper. For revealing the variables most predictive of youth prediabetes/diabetes, the EI model with the highest average BA was interpreted.

The code implementing all the prediction methods and their evaluation is available for download from POND.

**References**


1. The SAS Program for CDC Growth Charts, (available at https://www.cdc.gov/nccdphp/dnpao/growthcharts/resources/sas.htm).
2. BernardRosner - Childhood Blood Pressure Macro-batch mode, (available at https://sites.google.com/a/channing.harvard.edu/bernardrosner/pediatric-blood-press/childhood-blood-pressure).
3. United States Department of Agriculture (USDA) Food Consumption and Nutrient Intake, (available at https://www.ers.usda.gov/data-products/food-consumption-and-nutrient-intakes/food-consumption-and-nutrient-intakes/#Current%20Data%20Files).
4. G. Zipf, M. Chiappa, K. S. Porter, Y. Ostchega, B. G. Lewis, J. Dostal, National health and nutrition examination survey: plan and operations, 1999-2010. Vital Health Stat. Ser 1 Programs Collect. Proced., 1–37 (2013).
5. Ensemble Integration (EI): Integrating multimodal data through interpretable heterogeneous ensembles (2023), (available at https://github.com/GauravPandeyLab/ei-python).
6. R. Caruana, A. Munson, A. Niculescu-Mizil, "Getting the Most Out of Ensemble Selection" in Sixth International Conference on Data Mining (ICDM'06) (2006), pp. 828–833.
7. R. Caruana, A. Niculescu-Mizil, G. Crew, A. Ksikes, "Ensemble selection from libraries of models" in Proceedings of the twenty-first international conference on Machine learning (Association for Computing Machinery, New York, NY, USA, 2004; https://doi.org/10.1145/1015330.1015432), ICML '04, p. 18.
8. S. Arslanian, F. Bacha, M. Grey, M. D. Marcus, N. H. White, P. Zeitler, Evaluation and Management of Youth-Onset Type 2 Diabetes: A Position Statement by the American Diabetes Association. Diabetes Care. 41, 2648–2668 (2018).
9. K. H. Brodersen, C. S. Ong, K. E. Stephan, J. M. Buhmann, "The Balanced Accuracy and Its Posterior Distribution" in 2010 20th International Conference on Pattern Recognition (2010), pp. 3121–3124.
10. Y. Benjamini, Y. Hochberg, Controlling the False Discovery Rate: A Practical and Powerful Approach to Multiple Testing. J. R. Stat. Soc. Ser. B Methodol. 57, 289–300 (1995).
